# Risk factors for intensive care admission and death amongst children and young people admitted to hospital with COVID-19 and PIMS-TS in England during the first pandemic year

**DOI:** 10.1101/2021.07.01.21259785

**Authors:** J L Ward, R Harwood, C Smith, S Kenny, M Clark, PJ Davis, ES Draper, D Hargreaves, S Ladhani, M Linney, K Luyt, S Turner, E Whittaker, L K Fraser, R.M Viner

**Affiliations:** UCL Great Ormond St. Institute of Child Health, London; Institute of Systems, Molecular and Integrative Biology, University of Liverpool, Liverpool; NHS England and Improvement; Paediatric Intensive Care Unit, Bristol Royal Hospital for Children, Bristol; PICANet, Department of Health Sciences, University of Leicester, Leicester; Imperial College London, Department of Primary Care and Public Health; Immunisation and Countermeasures, Public Health England; University Hospitals Sussex NHS Foundation Trust; Bristol Medical School, University of Bristol, Bristol; Royal College of Paediatrics and Child Health; University of Aberdeen; Imperial College School of Medicine, London; Martin House Research Centre, Dept of Health Sciences, University of York

**Author notes:** **Correspondence**: Dr. Joseph Ward, UCL Great Ormond St. Institute of Child Health, 30 Guilford St. London WC1N 1EH.

## Abstract

Identifying which children and young people (CYP) are vulnerable to severe disease following SARS-CoV-2 is important to guide shielding and vaccination policy.

**Methods:** We used data for all inpatient hospital admissions in England in CYP aged 0-17 between March 1^st^ 2015 to Feb 28^th^ 2021, linked to paediatric intensive care unit (PICU), SARS-CoV-2 PCR testing, and mortality data. We examined associations between PICU admission and death by sociodemographic factors and comorbidities within COVID-19 and PIMS-TS admissions. We calculated odds ratios and predicted probability of PICU admission using generalized estimation equations, and compared these between COVID-19, PIMS-TS, other admissions in 2020/21, all admissions in 2019/20, and admissions due to influenza in 20219/20. Analyses of deaths were descriptive due to low numbers.

**Findings:** Within COVID-19, there were 6,338 hospital admissions, 259 PICU admissions and 8 deaths. Within PIMS-TS there were 712 hospital admissions 312 PICU admissions and <5 deaths. Males were 52.8% of COVID-19 admissions (similar to other causes of admission), but were 63.5% of PIMS-TS admissions. CYP aged 10-17 were 35.6 and 29% of COVID-19 and PIMS-TS admissions respectively, higher than in all admission and influenza admissions in 2019/20. In multivariable models, odds of PICU admission were: increased amongst neonates and decreased amongst 15-17 compared with 1-4 year olds with COVID-19, increased in older CYP and females with PIMS-TS, and increased for Black compared with White ethnicity in COVID-19 and PIMS-TS. Odds of PICU admission with COVID-19 were increased for CYP with any comorbidity and were highest for CYP with multiple medical problems. Increases in risk of PICU admission associated with comorbidities showed similar patterns for COVID-19 and all admissions in 2019/20 and influenza admissions in 2019/20, but were greater for COVID-19. Interpreting associations with comorbidities within PIMS-TS was complex due to the multisystem nature of the disease.

**Interpretation:** CYP were at very low risk of severe disease and death from COVID-19 or PIMS-TS. PICU admission due to PIMS-TS was associated with older non-white CYP. Patterns of vulnerability for severe COVID-19 appear to magnify background risk factors for serious illness in CYP.

**Evidence before this study:** We conducted a systematic review and meta-analysis of studies investigating risk factors associated with severe disease among children and young people admitted with COVID-19 and PIMS-TS, [Harwood, R et al. 2021 (submission to the Lancet linked with this paper)]. We identified 81 studies. Infants were found to have increased odds of PICU admission compared with 1-4, but there were no associations by sex. Other factors associated with PICU admission included number of co-morbid conditions, with neurological, cardiac and gastrointestinal associated with the greatest risk. Low numbers of serious SARS-CoV-2 infections or deaths amongst CYP limit these analyses, yet national studies of CYP have not yet been published. Importantly, we found these studies did not take into account background risks for severe illness in CYP who are known to be vulnerable before the pandemic.

**What this study adds:** This is the first population base study of risk factors for severe disease following SARS-CoV-2 infection in CYP in England. We analyse all admissions to hospital amongst 0-17 year olds nationally between 2015-2021 linked to multiple other health datasets. We explore how socioeconomic factors and co-morbidities are associated with Paediatric Intensive Care Unit (PICU) admission and death amongst CYP admitted with COVID-19 and PIMS-TS, and compare this with other causes for admission during the pandemic and in the year prior. As CYP with PIMS-TS are highly likely to require hospitalization, we were able to analyse total national cases of the condition during 2020/21.

We found extremely low numbers of CYP required PICU or died as a result of SARS-CoV-2 in the first pandemic year. CYP admitted due to COVID-19 disease were older and more likely to be non-white with pre-existing conditions, similar to patterns seen in adults. Patterns of associations between comorbidities and risk of PICU admission amongst COVID-19 were similar to those seen for all admissions and influenza admissions in the year prior to the pandemic. However, the increase in risk associated with comorbidities for COVID-19 admissions were greater than in these cohorts.

We found most cases of PIMS-TS were amongst non-white male adolescents without previous hospital admissions. Interpreting associations between comorbidities and PICU admission for PIMS-TS was complicated by the multi-system nature of the disease.

**Implications of all the available evidence:** CYP with most vulnerable to COVID-19 were also those most at risk of prior to the pandemic due to other illnesses such as influenza, although COVID-19 appears to amplify this risk profile. It is important to consider this context when advising parents and carers regarding the risk posed by COVID-19, considering potential harms to CYP as a result of shielding precautions.

## Background

Most children and young people (CYP) experience a mild disease following SARS-CoV-2 infection compared with adults,^1-3^ and asymptomatic infection is common.^4^ However, severe clinical outcomes have been reported amongst CYP due to COVID-19 and Paediatric Inflammatory Multisystem Syndrome Temporally Associated with SARS-CoV-2 (PIMS-TS), including a small number of deaths.^2,5-7^ Understanding which CYP are vulnerable to increased risk is important to guide clinicians, families and policymakers in relation to protective shielding and potential vaccination strategies.

Early in the pandemic, guidance from the UK Royal College of Paediatrics and Child Health (RCPCH) identified CYP with immunodeficiency or immunosuppression, and those with certain malignancies, as having greatest vulnerability to COVID-19.^8^ However, CYP with a broad range of other conditions have also been highlighted as being potentially clinically extremely vulnerable (CEV). CYP who are identified as CEV have been advised to take additional “shielding” precautions to reduce the risk of SARS-CoV-2 infection in many countries. These include measures which may result in potential harm to CYP and their families, including those associated with reduced social mixing and not returning to school.

Clear guidance is urgently needed on which CYP are at higher risk of poorer outcomes in order to limit harms due to inappropriate shielding. The rarity of severe and fatal COVID-19 in CYP means that large-scale population-based studies are needed to identify CYP at most at risk. Theses analyses also need to take into account background risks for severe illness that preceded the pandemic; CYP who are at increased risk of SARS-CoV-2 infection may also be those who are vulnerable to other respiratory viruses such as influenza.^9^

We used national linked administrative health data from England to examine risk factors for intensive care admission and death in CYP admitted to hospital with SARS-CoV-2 infection during the first pandemic year 2020-21, and compared this amongst CYP admitted with other causes of admission that year, and admissions during 2019-20 including those due to influenza.

## Methods

### Data

We used Secondary Use Services (SUS), an administrative national database covering ∼98% of National Health Service (NHS) hospital activity in England.^10^ Data were available for admissions due to any cause in CYP aged 0-17 years in England between March 1^st^ 2015 to Feb 28^th^ 2021. (n=11,467,027). Coded fields included for reason for hospital admission, co-morbidities, and sociodemographic characteristics. However, as clinical details are limited in SUS, and to aid identification of COVID-19 and PIMS-TS admissions, these data were deterministically linked using unique patient NHS numbers to the following healthcare datasets:

1. Paediatric Intensive Care Audit Network (PICANet) data containing all Paediatric Intensive Care (PICU) admissions in England.
2. Death registrations provided by the Office for National Statistics (ONS).^11,12^
3. National Child Mortality Database (NCMD), which collects preliminary notification data within 48 hours of death of a CYP death in England and Wales
4. SARS-CoV-2 PCR-testing data provided by Public Health England (PHE).

## Outcomes and Exposures

We examined associations between severity outcomes (PICU admission or death) and the following exposures: reason for hospital admission (COVID-19, PIMS-TS, other), sociodemographic factors and presence of comorbidities.

We linked hospitalisations in SUS to PICANet data if the PICU admission date occurred during or within one day of the SUS admission or discharge date, to account for coding error in either dataset. We defined admissions resulting in death as the last admission for each CYP that occur within 28 days of death identified through ONS or NCMD. All NCMD deaths during the pandemic were clinically reviewed as part of a separate analysis [Smith, C et al 2021 (submission to the Lancet linked with this paper)] to identify the contribution of SARS-CoV-2 infection and PIMS-TS to death.

### Reason for admission

We used primary and secondary diagnoses coded to International Classification of Diseases 10 (ICD 10) to define all hospital admissions between 1^st^ Feb 2019 and 31^st^ Jan 2021. We excluded all traumatic admissions and non-emergency admissions (i.e. elective or maternity/newborn) from the analysis, and classified the remainder into five cohorts:

- admissions due to COVID-19 (1^st^ Feb 2020 – 31^st^ Jan 2021);
- admissions due to PIMS-TS (1^st^ Feb 2020 – 31^st^ Jan 2021);
- all other admissions during the pandemic year (1^st^ Feb 2020 – 31^st^ Jan 2021);
- all admissions in the year prior to the pandemic (1^st^ Feb 2019 – 31^st^ Jan 2020);
- all admissions where the primary diagnosis was influenza in the year prior to the pandemic (1^st^ Feb 2019 – 31^st^ Jan 2020).

As quality and completeness of coding within SUS is variable, we used the whole SUS dataset (2015-2021) to populate socio-demographic and comorbidity data for CYP from all types of admission (emergency, elective and maternal).

We defined COVID-19 admissions as those occurring after Feb 1^st^ 2020 with relevant ICD-10 codes recorded as reason for admission, or (using linked data) where there was a positive PCR test for SARS-CoV-2 within 7 days of admission or discharge, (unless this occurred at least 7 days after admission to PICU, and nosocomial infection was likely).

We defined PIMS-TS admissions those occurring after 1^st^ Feb 2020 with ICD-10 codes recorded reason for admission for either PIMS-TS (introduced November 2020), or Kawasaki disease or systemic inflammatory response syndrome, (used as proxies for PIMS-TS prior to November 2020).

To improve the identification of COVID-19 and PIMS-TS, we also reviewed details of all PICU admissions during the pandemic held within PICANet. Where treating specialists determined the PICU admission was due to either COVID-19 or PIMS-TS, we recoded the SUS admission accordingly. Hospital admissions identified as both due to COVID-19 and PIMS-TS were defined as being due to PIMS-TS, as we assumed the COVID-19 diagnosis was part of the same disease process.

### Socio-demographic exposures

Age was categorised as neonates (admission within 1 month of birth), post-natal infants (admission between 1 – 12 months of birth), 1-4 years, 5-9 years, 10-14 years and 15-17 years. We defined ethnicity as: White, Mixed, Asian, Black, Other and unknown. We used Index of Multiple Deprivation (IMD) 2019 quintile category (hereafter IMD category) to define area level socioeconomic status of CYP.

### Co-morbidities

We used published literature and guidance on shielding to identify co-morbidities likely to increase risk of severe SARS-CoV-2 disease.^8^ [Harwood, R et al. 2021 (submission to the Lancet linked with this paper)]. We identified CYP with chronic medical conditions by body system, those with life-limiting conditions, and those with asthma, diabetes, epilepsy, sickle cell disease and trisomy 21, using recognised ICD-10 code lists.^13,14^ We also identified CYP with multiple medical problems as those with: comorbidities in more than one body system; comorbidities in both neurological and respiratory, neurological and cardiovascular, or respiratory and cardiovascular body systems. We compared the presence of each comorbidity in CYP to those with no co-morbidities recorded in all analyses.

## Analyses

First we described the characteristics of each of the five cohorts: admissions due to COVID-19, admissions due to PIMS-TS, other non-traumatic admissions in 2020/21 (hereafter “other pandemic year admissions”), all non-traumatic admissions in 2019/20, and admissions due to influenza in 2019/20.

We then modelled the association between sociodemographic factors and co-morbidities with PICU admission within each cohort separately. All analyses were performed in Stata 16 (StataCorp, College Station TX). Models employed generalized estimation equations (GEE) using the *xtgee* command in order to account for multiple admissions within the same CYP across and within different cohorts. Models used a logit link, specifying the covariance structure as “exchangeable” (i.e. assumed equal correlations between any two admissions within one CYP). We then calculated the difference in predicted probability for PICU admission amongst those with and without each comorbidity using the *margins* post estimation command. We ran univariable and then multi-variable models adjusting for age, sex, ethnic group and IMD category within each cohort. Comparisons between cohorts were not tested; significance was inferred if 95% confidence intervals did not overlap. We were unable to model death as an outcome in these analyses due to low numbers. In sensitivity analyses, we repeated analyses to only include secondary school age CYP to inform vaccination policy (i.e. ages 11-17).

### Ethics approval and legal basis for data linkage and analyses

Ethics approval was provided after review by Yorkshire and the Humber, South Yorkshire NHS Research Ethics Committee on 10^th^ June 2021 (Reference 21/YH/0127).

Current Control Of Patient Information (COPI) regulations provide a legal basis for linking these datasets without consent.^15^

### NCMD

The NCMD legal basis to collect confidential and personal level data under the Common Law Duty of Confidentiality has been established through the Children Act 2004 Sections M - N, Working Together to Safeguard Children 2018 (https://consult.education.gov.uk/child-protection-safeguarding-and-family-law/working-together-to-safeguard-children-revisions-t/supporting_documents/Working%20Together%20to%20Safeguard%20Children.pdf) and associated Child Death Review Statutory & Operational Guidance https://assets.publishing.service.gov.uk/government/uploads/system/uploads/attachment_data/file/859302/child-death-review-statutory-and-operational-guidance-england.pdf). The NCMD legal basis to collect personal data under the General Data Protection Regulation (GDPR) without consent is defined by GDPR Article 6 (e) Public task and 9 (h) Health or social care (with a basis in law).

### PICANet

Processing of personally identifiable data for the purposes of service evaluation, audit, and research was approved by the Patient Information Advisory Group (now the Health Research Authority Confidentiality Advisory Group) in 2002 under Section 60 of the Health and Social Care Act (subsequently Section 251 of the National Health Service Act 2006) (reference: PIAG 4-07(c) 2002). This was amended and approved specifically to collect additional data relating to COVID-19 for confirmed and suspected cases.

## Results

There were 1,242,197 emergency non-traumatic admissions to secondary care in England (hereafter “admissions”) between 01 Feb 2019 and 31 Jan 2021 involving 892,906 CYP; 699,397 (78%) had only one admission. During 2020/21, there were 470,606 admissions: 6,338 with COVID-19 amongst 5,830 CYP; 712 with PIMS-TS amongst 690 CYP and 463,556 for other causes amongst 367,637 CYP. This compared with 771,591 admissions for 587,115 CYP during 2019/20, of which 6968 were due to influenza in 6,780 CYP (see supplementary material 1 table S1 and S15). 69.8% of CYP admitted due to PIMS-TS had no prior hospital admissions, compared with 49.5-54.4% in all other cohorts.

The distribution of admissions by age, sex and ethnicity differed between the COVID-19, PIMS-TS, and other cohorts (Table 1). A higher proportion of the PIMS-TS cohort was male (63.5%) compared to the other cohorts (52.8-54%). Overall, 30.9% of admissions with COVID-19 were in infants, similar to other pandemic year admissions and total admissions in 2019/20, but more than for influenza (17.1%). Amongst PIMS-TS, only 10.3% of admissions were in infants, where >85% were among 1-14 year-olds. CYP with non-White ethnicity made up 41.9% of COVID-19 admissions, and 60.0% of PIMS-TS admissions, higher than the other cohorts. There were more admissions in CYP from the most deprived IMD category compared with the least deprived in all cohorts.

**Table 1.**
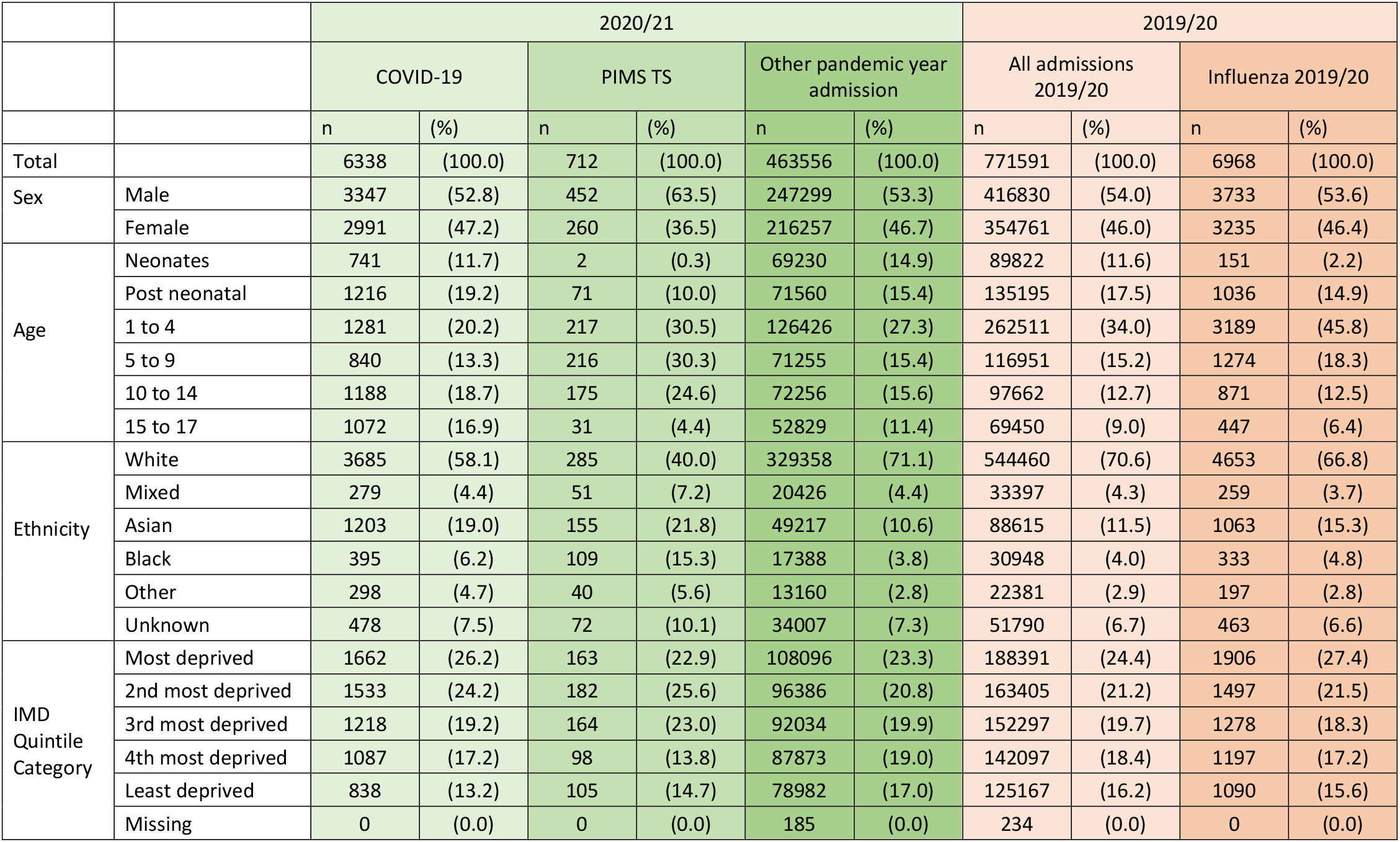
Number and proportion of admissions by sociodemographic characteristics within each cohort (COVID-19, PIMS-TS, other pandemic year admissions; all admissions in 2019/20; influenza admissions in 2019/20)

|  |  | 2020/21 |  |  |  |  |  | 2019/20 |  |  |  |
| --- | --- | --- | --- | --- | --- | --- | --- | --- | --- | --- | --- |
|  |  | COVID-19 |  | PIMS TS |  | Other pandemic year admission |  | All admissions 2019/20 |  | Influenza 2019/20 |  |
|  |  | n | (%) | n | (%) | n | (%) | n | (%) | n | (%) |
| Total |  | 6338 | (100.0) | 712 | (100.0) | 463556 | (100.0) | 771591 | (100.0) | 6968 | (100.0) |
| Sex | Male | 3347 | (52.8) | 452 | (63.5) | 247299 | (53.3) | 416830 | (54.0) | 3733 | (53.6) |
|  | Female | 2991 | (47.2) | 260 | (36.5) | 216257 | (46.7) | 354761 | (46.0) | 3235 | (46.4) |
| Age | Neonates | 741 | (11.7) | 2 | (0.3) | 69230 | (14.9) | 89822 | (11.6) | 151 | (2.2) |
|  | Post neonatal | 1216 | (19.2) | 71 | (10.0) | 71560 | (15.4) | 135195 | (17.5) | 1036 | (14.9) |
|  | 1 to 4 | 1281 | (20.2) | 217 | (30.5) | 126426 | (27.3) | 262511 | (34.0) | 3189 | (45.8) |
|  | 5 to 9 | 840 | (13.3) | 216 | (30.3) | 71255 | (15.4) | 116951 | (15.2) | 1274 | (18.3) |
|  | 10 to 14 | 1188 | (18.7) | 175 | (24.6) | 72256 | (15.6) | 97662 | (12.7) | 871 | (12.5) |
|  | 15 to 17 | 1072 | (16.9) | 31 | (4.4) | 52829 | (11.4) | 69450 | (9.0) | 447 | (6.4) |
| Ethnicity | White | 3685 | (58.1) | 285 | (40.0) | 329358 | (71.1) | 544460 | (70.6) | 4653 | (66.8) |
|  | Mixed | 279 | (4.4) | 51 | (7.2) | 20426 | (4.4) | 33397 | (4.3) | 259 | (3.7) |
|  | Asian | 1203 | (19.0) | 155 | (21.8) | 49217 | (10.6) | 88615 | (11.5) | 1063 | (15.3) |
|  | Black | 395 | (6.2) | 109 | (15.3) | 17388 | (3.8) | 30948 | (4.0) | 333 | (4.8) |
|  | Other | 298 | (4.7) | 40 | (5.6) | 13160 | (2.8) | 22381 | (2.9) | 197 | (2.8) |
|  | Unknown | 478 | (7.5) | 72 | (10.1) | 34007 | (7.3) | 51790 | (6.7) | 463 | (6.6) |
| IMD Quintile Category | Most deprived | 1662 | (26.2) | 163 | (22.9) | 108096 | (23.3) | 188391 | (24.4) | 1906 | (27.4) |
|  | 2nd most deprived | 1533 | (24.2) | 182 | (25.6) | 96386 | (20.8) | 163405 | (21.2) | 1497 | (21.5) |
|  | 3rd most deprived | 1218 | (19.2) | 164 | (23.0) | 92034 | (19.9) | 152297 | (19.7) | 1278 | (18.3) |
|  | 4th most deprived | 1087 | (17.2) | 98 | (13.8) | 87873 | (19.0) | 142097 | (18.4) | 1197 | (17.2) |
|  | Least deprived | 838 | (13.2) | 105 | (14.7) | 78982 | (17.0) | 125167 | (16.2) | 1090 | (15.6) |
|  | Missing | 0 | (0.0) | 0 | (0.0) | 185 | (0.0) | 234 | (0.0) | 0 | (0.0) |

Amongst COVID-19 admissions, 53.9% had a recorded comorbidity, and 18.0% had a life limiting comorbidity, higher than for other pandemic year admissions, all admissions in 2019/20, and influenza admissions in 2019/20 (Supplementary Table S1). Patterns of comorbidities amongst admissions with PIMS-TS were different to the other cohorts, with 68.3% having any comorbidity recorded, of which 20.6% were life-limiting. Due to the multi-system nature of PIMS-TS, many of the comorbidities recorded could have been related to complications of the disease, rather than prior conditions. Although 40.2% of PIMS-TS admissions had a cardiovascular comorbidity recorded, only 5.3% had a congenital cardiac condition, with remaining codes including arrhythmias and aneurysms which may reflect the disease process. When non-congenital cardiac conditions, blood disorders and anaemias were excluded, only 15.9% of PIMS-TS admissions had a comorbidity recorded, compared with 30-35% in the other cohorts.

### Outcomes following admission

Table 2 shows total numbers and proportions of PICU admissions within each cohort by comorbidity category, with additional data for all comorbidities examined in Supplementary material 1 Tables S3-S5. Across COVID-19 admissions, 259 (4.1%) were admitted to PICU, compared with 312 (43.8%) of PIMS-TS admissions, 5016 (1.1%) of other pandemic year admissions, 7282 (0.9%) of all admissions in 2019/20 and 161 (2.3%) of influenza admissions in 2019/20.

**Table 2.**
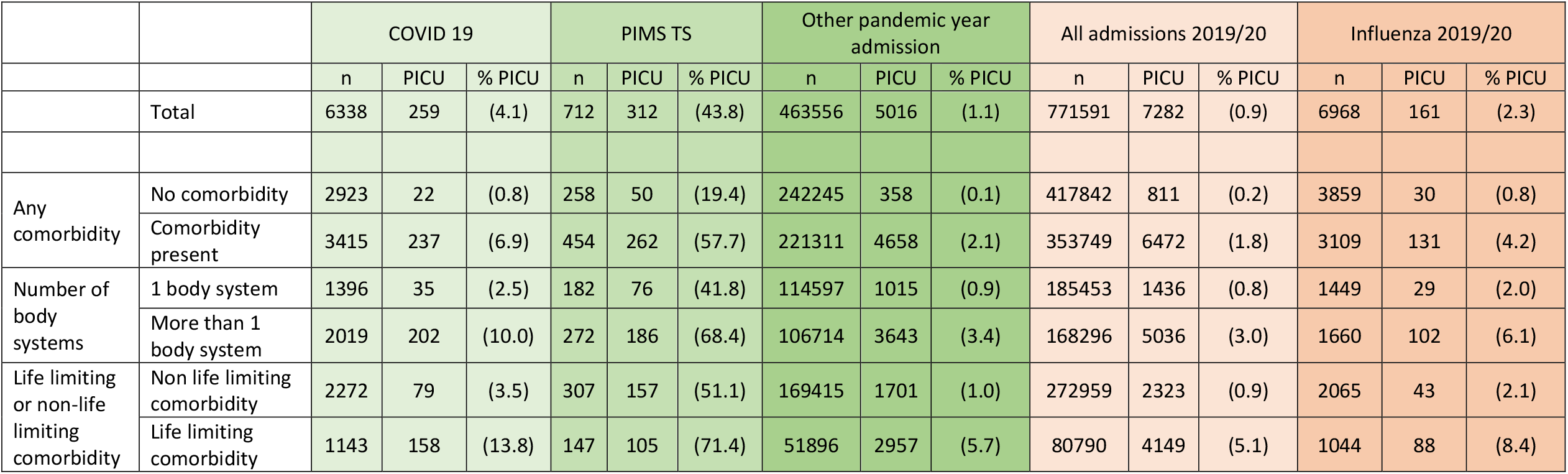
Total admissions and proportion resulting in paediatric critical care admission (PICU) by selected comorbidity groups within each cohort (COVID-19, PIMS-TS, other pandemic year admissions; all admissions in 2019/20; influenza admissions in 2019/20)

Twenty nine CYP admitted with COVID-19 died within 28 days of hospitalisation, of which 8 were confirmed as likely caused by SARS-CoV-2 infection; all had a comorbidity recorded and 7/8 had a life-limiting condition. Six CYP died within 28 days of an admission with PIMS-TS, of which < 5 were thought to be caused by the disease.

### Sociodemographic factors

In multivariable models adjusting for all factors and the presence of comorbidities, female sex was associated with increased odds of PICU admission for PIMS-TS, and reduced odds amongst all admissions 2019/20, with no associations found by sex for COVID-19 or the other cohorts (supplementary material 1, tables S11-S15). Compared with admissions amongst 1-4 year olds, odds of PICU admission for COVID-19 were increased amongst neonates and decreased amongst 15-17 year olds, similar to patterns for other pandemic year admissions and all admissions 2019/20, (although odds were also decreased for 5-14 year olds in these cohorts). Odds of PICU admission for PIMS-TS increased with age in a stepwise fashion and were highest in 15-17 year olds. The odds of PICU admission within influenza admissions in 2019/20 were only higher amongst neonates compared with 1-4 year olds.

Compared with White CYP, odds of PICU admission were higher amongst Black CYP for COVID-19 and Black, Asian and CYP with unknown ethnicity for PIMS-TS. Other pandemic year admissions and all admissions 2019/20 showed a pattern of higher odds of PICU admission in non-White ethnic groups, with no evident differences by ethnicity amongst influenza admissions in 2019/20. There were no significant differences in odds of PICU admission by IMD category for COVID-19, all admissions in 2019/20 and influenza admissions in 2019/20. In contrast, odds of PICU admission were decreased in less deprived categories amongst PIMS-TS admissions, and increased in less deprived categories amongst other pandemic year admissions.

### Comorbidities

The odds of admission to PICU were increased amongst CYP with any comorbidity compared with no comorbidity in all cohorts (supplementary material 2). The increases in odds of PICU admission associated with having each of any comorbidity, a life limiting comorbidity, or comorbidities in more than one body system for COVID-19 (Figure 1), had overlapping confidence intervals with those for all admissions in 2019/20 and influenza admissions in 2019/20, but were lower than for other pandemic year admissions. Odds ratios for PIMS-TS admissions were consistently the lowest of any cohort for each comorbidity category, although confidence intervals often overlapped.

**Figure 1.**
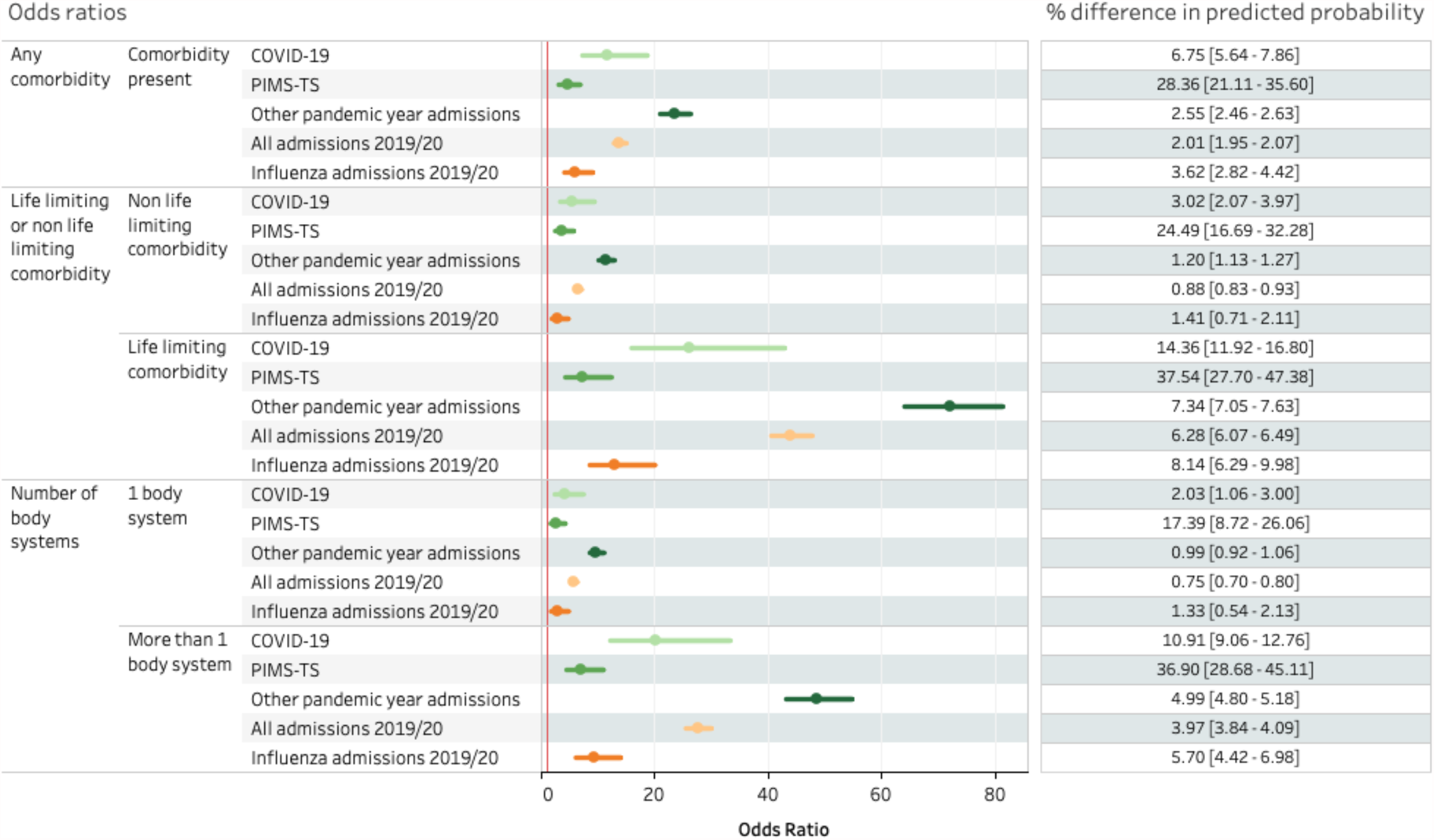
Odds ratios and percentage point difference in predicted probability with 95% confidence intervals for admission to PICU by comorbidity groups within each cohort, adjusted for age, sex, IMD category, ethnicity Notes: Results compare odds of PICU admission in selected comorbidity compared with no comorbidity. The red line indicates odds ratio of 1. Percentage point difference in predicted probability of PICU admission are shown for selected comorbidities compared with no comorbidity

For body system comorbidities (Figure 2), odds ratios for the increase associated with cancer/haem, neurological, respiratory, neurological with respiratory and respiratory with cardiovascular comorbidities in COVID-19 appeared comparable to influenza and all admissions 2019-20 but not PIMS-TS (where the increase in odds was lower) or other pandemic year admissions (where the increase in odds was higher). The increase in odds for cardiovascular comorbidities within COVID-19 appeared similar to that seen in all admissions in 2019/20, but higher than influenza admissions 2019/20 and PIMS-TS, and lower than for other pandemic year admissions. A similar pattern was observed for combinations of body-system comorbidities (Figure 3), i.e. that the increase in odds for COVID-19 appeared similar to that for influenza and all admissions 2019/20, but was higher than for PIMS-TS and lower than in other pandemic year admissions.

**Figure 2.**
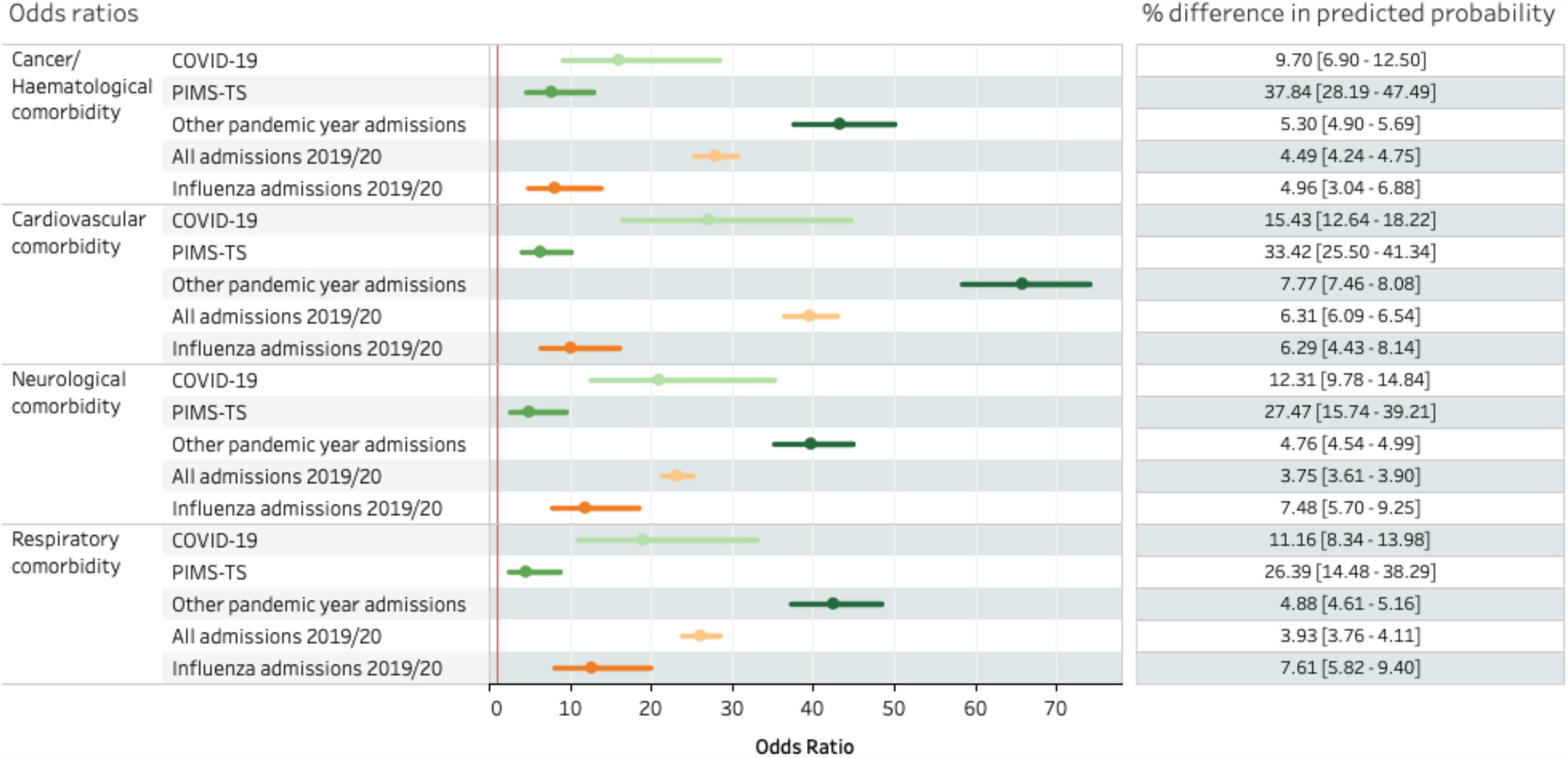
Odds ratios and percentage point difference in predicted probability with 95% confidence intervals for admission to PICU by body system comorbidities within each cohort, adjusted for age, sex, IMD category, ethnicity Notes: Results compare odds of PICU admission in selected comorbidity compared with no comorbidity. The red line indicates odds ratio of 1. Percentage point difference in predicted probability of PICU admission are shown for selected comorbidities compared with no comorbidity

**Figure 3.**
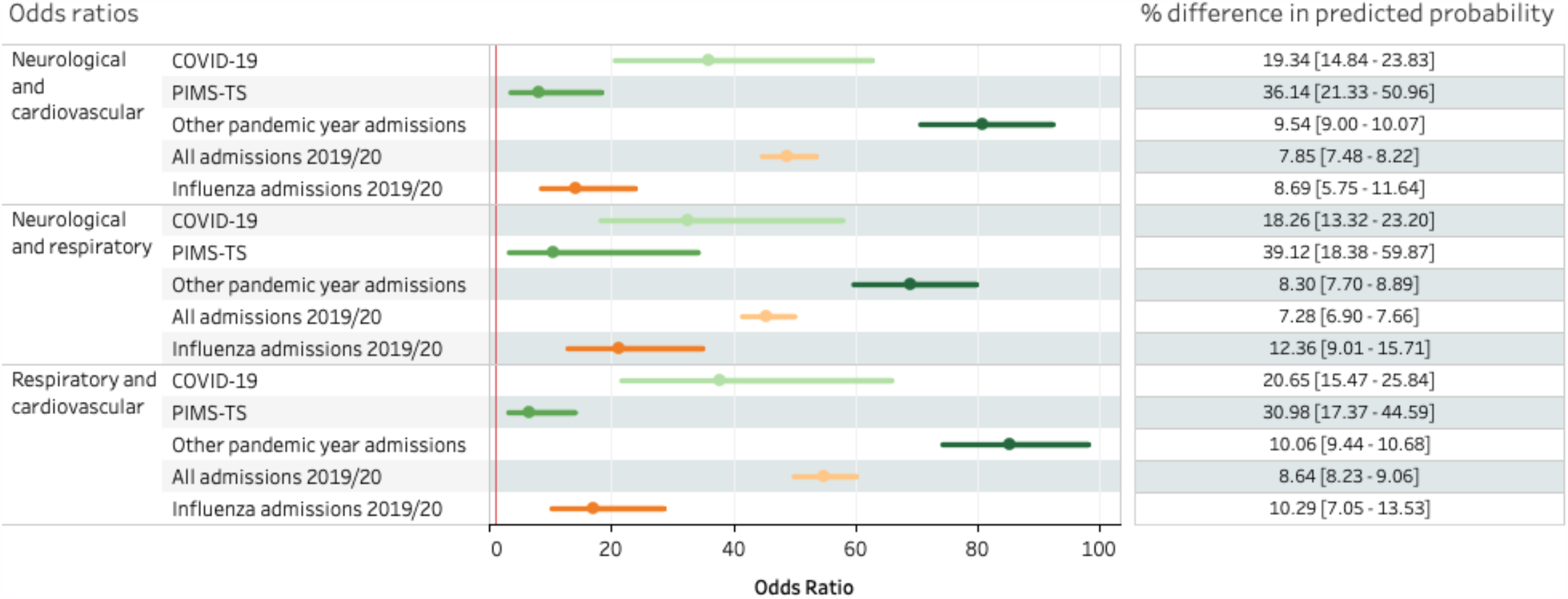
Odds ratios and predicted probability with 95% confidence intervals for admission to PICU by comorbidity combinations within each cohort, adjusted for age, sex, IMD category, ethnicity Notes: Results compare odds of PICU admission in selected comorbidity compared with no comorbidity. The red line indicates odds ratio of 1. Percentage point difference in predicted probability of PICU admission are shown for selected comorbidities compared with no comorbidity

Asthma, diabetes, epilepsy and trisomy 21 each increased risk of PICU admission for COVID-19, although sickle cell disease did not (Figure 4). Increases in odds for COVID-19 appeared broadly similar to those for other cohorts although confidence intervals were wide, particularly for PIMS-TS.

**Figure 4.**
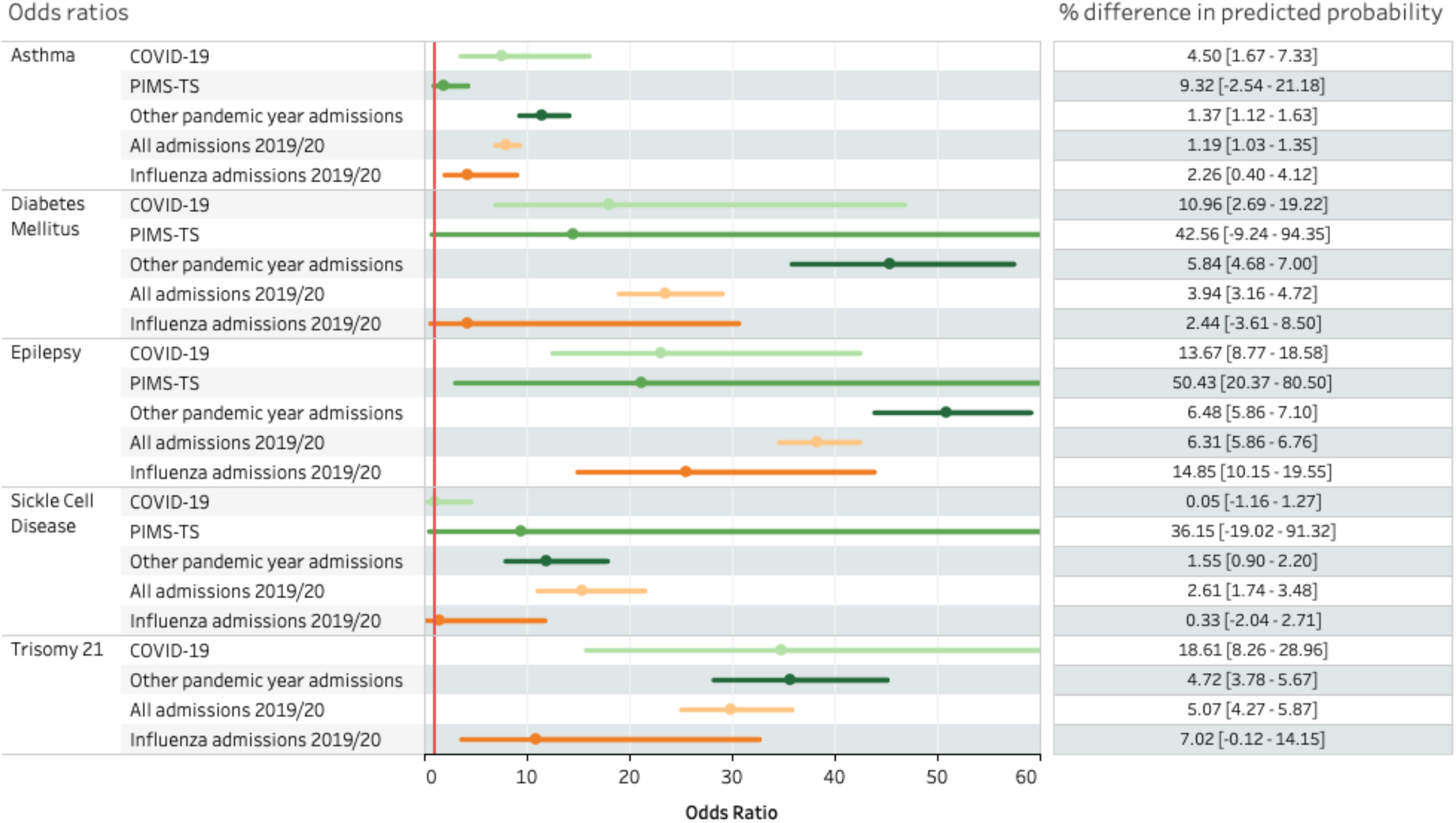
Odds ratios and predicted probability with 95% confidence intervals for admission to PICU by selected diagnoses within each cohort, adjusted for age, sex, IMD category, ethnicity Notes: Results compare odds of PICU admission in selected comorbidity compared with no comorbidity. Upper limits for confidence intervals have been truncated for selected comorbidities for clarity. The red line indicates odds ratio of 1. Percentage point difference in predicted probability of PICU admission are shown for selected comorbidities compared with no comorbidity.

Results from sensitivity analyses are shown in supplementary material 1 figures S1-S4 and supplementary material 3. Patterns of odds ratios were similar when only 11-17 year olds were included, although female sex was associated with significantly reduced odds of PICU admission for COVID-19. Increases in odds of PICU admission associated with comorbidities for COVID-19 amongst 11-17 were lower than when all CYP were included for some outcomes. However, due to low numbers, confidence intervals around these estimates were wide. We were not able to model associations within Influenza admissions in 11-17 due to low numbers.

## Discussion

We found very few CYP admitted to hospital in England due to COVID-19 or PIMS-TS went on to develop severe disease or die. Of the 12.02 million 0-17 year olds in England during 2020, 1 in 2062 (n= 5830) were admitted to hospital due to COVID-19, and 1 in 47,903 (n=251) were admitted to PICU. This represents only 1.3% of all secondary care admissions in the pandemic year and less than 5% of non-traumatic emergency PICU admissions. Eight of these CYP died. For PIMS-TS, 1 in 17,425 (n=690) of CYP in England were admitted to hospital, 1 in 38,911 (n=309) were admitted to PICU, and < 5 children died. This likely represents all PIMS-TS cases nationally over the study period, as the vast majority will have required hospitalisation.

CYP admitted with COVID-19 and PIMS-TS were older and more likely to be non-white than in the other cohorts examined. For COVID-19, we found the odds of PICU admission increased amongst neonates compared with 1-4 year-olds, and those who were Black compared with White ethnicity, but found no associations by deprivation. Female sex was associated with significantly lower olds of PICU admission for COVID-19, but only when in sensitivities where data were restricted to 11-17 year olds. For PIMS-TS, the odds of PICU admission were increased amongst females, older CYP and those from non-White ethnic groups.

Of the 251 CYP admitted to PICU with COVID-19, 91% (n=229) had an underlying condition or comorbidity. The odds of PICU admission due to COVID-19 were increased in all comorbidity categories tested except sickle cell disease. We found that CYP with complex medical problems across multiple body systems, and those with neurodisability, were at greatest risk. This pattern is described in previous work,^16^ and is consistent with our meta-analysis of the published data, where each increase in number of pre-existing conditions was associated with increased odds of PICU admission and death for COVID-19. Increases in odds of PICU admission associated with comorbidities in PIMS-TS were lower than for COVID-19, but are difficult to interpret; coding of PIMS-TS admissions suggested two-thirds had a comorbidity, whilst three quarters had no prior admissions to hospital. When codes which include known cardiac and haematological complications of PIMS-TS were excluded, estimates for comorbidities in these admissions dropped to around 15%.^17^

Our comparison with other causes of admission allowed us to assess whether these risk factors are specific to COVID-19 or PIMS-TS, or reflect background vulnerability to serious illness. Our findings that Asian and Black ethnicity was associated with increased odds of serious disease was similar to findings from other cohorts except for influenza. However, a high proportion of admissions for COVID-19 and PIMS-TS were from non-White ethnic groups, consistent with previous work,^18-20^ and increases in odds associated with non-White ethnicity were greater in these cohorts, similar to findings in adults.^21,22^ Age-patterns for COVID-19, and particularly for PIMS-TS admission, were notably shifted towards older age-groups in comparison with other cohorts, including influenza. We only found significant sex differences in risk for COVID-19 amongst 11-17 year olds, unlike other pandemic-year admissions and all admissions 2019/20, where female sex was associated with lower odds of PICU admission in all models. Almost two thirds of PIMS-TS admissions were amongst males, higher than in all other cohorts, but odds of PICU admission were greater amongst females.

We found broadly similar increases in odds for PICU admission associated with number of body systems or type of comorbidities across COVID-19, all 2019-20 admissions and influenza admissions. Increases in odds were highest for combinations of body system comorbidities e.g. neurological and respiratory, neurological and cardiovascular and respiratory and cardiovascular. Similarly, for the specific conditions examined, odds ratios overlapped with those for other pandemic year, all admissions 2019-20 and influenza, with the exception of sickle cell disease which was not associated with an increased odds of PICU admission for COVID-19 or influenza.

When absolute risk was examined, the increases in risk associated with comorbidities were relatively small in the COVID-19, other pandemic year, all admissions 2019-20 and influenza cohorts, although greater for COVID-19 than other groups. For example, for the 229 CYP with comorbidities admitted to PICU with COVID-19, the increase in risk above those without comorbidities was 2% for COVID-19, 0.75% for all admissions in 2019-20 and 1.3% for influenza. Combinations of comorbidities increased risk the most, although again numbers were very small. Amongst the 414 admissions with respiratory and neurological comorbidities, the increase in risks were 18.6% for COVID-19 compared with 12.3% for influenza and 7-8% for other cohorts. Whilst this greater increase in absolute risk with COVID-19 appeared significant for body system comorbidities and their combinations, confidence intervals overlapped for all specific conditions.

Our finding that the pattern of risks for severe COVID-19 related to comorbidities is similar to that for other reasons for admission suggests these reflect underlying vulnerabilities to illness and infection. A similar observation has been made in adults when risks were examined across COVID-19 and non-COVID deaths during the pandemic.^23^ However, whilst the pattern of risks was very similar and absolute risks remained relatively small, increases in absolute risk of PICU admission were often higher for COVID-19 than for other cohorts including influenza. This suggests that SARS-CoV-2 infection may magnify underlying risks faced by CYP with chronic and life-threatening conditions. It is also possible that these findings reflect changes in health system factors during the pandemic, although other studies have suggested there was no overall change in thresholds for PICU admission in England.^24^

Patterns within admissions due to COVID-19 amongst CYP, (older age, non-White ethnicity and presence of comorbidities), are very similar to those identified for adults.^21,22^ This suggests that the strong age-related risk of severe disease in adult COVID-19^22,25^ extends across the early life-course, but has previously been difficult to elicit in CYP due to the extreme rarity of severe disease.

### Limitations

Our study is subject to a number of limitations. We are unable to account for the effect of protective shielding on differential exposure to SARS-Cov-2 among CYP thought to be vulnerable, which may have affected our estimates. However, our findings relate to risk factors for severe disease once hospitalised, whereas shielding is likely to bias estimates of risk factors for infection, which we did not examine.

Missing or inaccurate data fields within HES or other datasets, and incomplete data linkage, may have affected our findings. We included both cause of admission and PCR testing for SARS-Cov-2 to identify CYP with COVID-19 to ensure we capture all likely cases, but this will have affected our case definition specificity. Identifying PIMS-TS cases was particularly problematic, as ICD-10 codes for this condition were only introduced several months into the pandemic. We included CYP coded with Kawasaki disease and systemic inflammatory response syndrome when examining PIMS-TS, some of whom will not have had the disease. We were unable to fully distinguish between admissions *with* COVID-19 and those *due to* COVID-19, and some of the admissions we classify as COVID-19 will include incidental positive PCR tests. We defined severe disease as CYP who were admitted to PICU, and were unable to assess the level of intensive support required. Our results may also have been affected by changes to thresholds for PICU admission, and coding practices, as the pandemic progressed. Our estimate for number of deaths due to COVID-19 and PIMS-TS only include hospitalised CYP. However, our separate study of all CYP deaths recorded in the pandemic year will allow for a more complete analysis of mortality risk associated with SARS-CoV-2 [Smith, C et al 2021 (submission to the Lancet linked with this paper)].

We use ICD-10 codes developed to identify chronic conditions across five years of admission data, and may have missed diagnoses recorded prior to this. Further, the ICD-10 codes we used included diagnoses which may relate to complications of acute disease, rather than pre-existing conditions only, as highlighted with PIMS-TS. We were unable to only include comorbidities prior to the index case to investigate this further as many CYP had no prior records, and this approach would not account for incomplete coding in previous admissions or diagnoses made in primary care. Linking HES data with national primary care records would improve identification of pre-existing conditions for these analyses, but these data are currently not available. Our analysis of individual or body system comorbidities does not account for CYP with both the comorbidity of interest and other conditions. However, we do assess odds of PICU by number of body systems involved, which does address identifying CYP with multiple medical problems. Finally, we were unable to examine some important risk factors for severe disease in adults in these analyses, including obesity,^26^ due to incomplete coding for these data.

## Conclusions

In marked contrast to adults, CYP were at very low risk of severe disease and death from COVID-19 or PIMS-TS during the first pandemic year. In the rare instances when CYP did require hospitalisation, risk factors for severe disease were similar to those reported for adults. Additionally, the pattern of comorbidities was similar to that seen with influenza and all admissions 2019/20, reflecting underlying vulnerabilities to infection, although COVID-19 magnified these risks to a small degree. We identified important demographic factors which were associated with PICU admission due to PIMS-TS. Further work is needed to understand the interaction of risk factors for infection and for severe disease in both COVID-19 and PIMS-TS.

## Data Availability

Some data are available on application to the authors

