## Supplement 1 for "Risk factors for intensive care admission and death amongst children and young people admitted to hospital with COVID-19 and PIMS-TS in England during the first pandemic year"

Supplementary Material 1

**Supplementary Tables**

**Supplementary figures**

### Supplementary Methods

#### Identifying COVID-19 and PIMS-TS admissions

We used guidance from NHS Digital to classify admissions due to COVID-19 and PIMS-TS using ICD-10 codes introduced during the pandemic.^13^ ICD-10 codes for COVID-19 (U071, U072) were introduced for use from February 2020, with further codes added in November 2020 (U073 U074), in addition to a specific code for PIMS-TS (U075). Prior to November 2020, PIMS-TS had been primarily coded within SUS using M303 (Kawasaki disease) and R65 (systemic inflammatory response syndrome) and identifying cases early in the pandemic was problematic. We defined COVID-19 admissions as those occurring after Feb 1^st^ 2020 with U071, U072, U073 or U074 recorded as either primary or secondary diagnosis, or (using linked data) where there was a positive PCR test for SARS-CoV-2 within 7 days of the admission or discharge date. Admissions where the first positive SARS-CoV-2 test occurred at least 7 days after admission to PICU were excluded, as these likely represent nosocomial infection in PICU, rather than the reason for intensive care.

We defined PIMS-TS admissions as those occurring after 1^st^ Feb 2020 with R65, M303 or U075 as a primary diagnosis only, as we were unable to distinguish between secondary diagnoses related to current or historic admissions with R65 and M303. To improve the identification of COVID-19 and PIMS-TS we also reviewed details of all PICU admissions during the pandemic held within PICANet. Where the PICANet data showed that treating specialists determined the PICU admission was due to either COVID-19 or PIMS-TS, we recoded the SUS admission accordingly. Hospital admissions identified as both due to COVID-19 and PIMS-TS were defined as being due to PIMS-TS, as we assumed the COVID-19 diagnosis was part of the same disease process.

#### Defining Socioeconomic status

We used Index of Multiple Deprivation (IMD) 2019 to define area level socioeconomic status of CYP. IMD is a measure of relative deprivation by Lower Super Output Area (LSOA), a small geographic area with a population of between 1000 to 3000 people (mean 1500) and containing between 400 and 1200 households.^14^ We calculated IMD quintile category (hereafter IMD category), weighted by total population of 0-17 year olds in 2019 within each LSOA to account for the increased child population size of those from deprived areas. Some LSOAs in SUS data referred to the 2001 census and were not identified in IMD 2019, which uses LSOAs from the 2011 census (n=810). We mapped these to LSOAs which appear in the 2011 census using data provided by ONS and available at data.gov.uk,^15^ taking the first value ordered alphabetically where LSOAs had been partitioned. We then assigned IMD category using the most recent value for LSOA across all admissions within each CYP.

| **Table S1** Number and proportion of admissions by comorbidity category within each cohort (COVID-19, PIMS-TS, other pandemic year admissions; all admissions in 2019/20; influenza admissions 2019/20) | | | | | | | | | | | |
| --- | --- | --- | --- | --- | --- | --- | --- | --- | --- | --- | --- |
|  |  | 2020/21 | | | | | | 2019/20 | | | |
|  |  | COVID 19 | | PIMS TS | | Other pandemic year admission | | All admissions 2019/20 | | Influenza 2019/20 | |
|  |  | n | (%) | n | (%) | n | (%) | n | (%) | n | (%) |
| Any comorbidity | No comorbidity | 2923 | (46.1) | 258 | (36.2) | 242245 | (52.3) | 417842 | (54.2) | 3859 | (55.4) |
|  | Comorbidity present | 3415 | (53.9) | 454 | (63.8) | 221311 | (47.7) | 353749 | (45.8) | 3109 | (44.6) |
| Any comorbidity* *excluding non-congenital cardiac conditions, anaemias and blood disorders | No comorbidity | 4205 | (66.4) | 599 | (84.1) | 299340 | (64.6) | 505475 | (65.5) | 4849 | (69.6) |
|  | Comorbidity present | 2133 | (33.6) | 113 | (15.9) | 164216 | (35.4) | 266116 | (34.5) | 2119 | (30.4) |
| Number of body systems | 1 body system | 1396 | (22.0) | 182 | (25.6) | 114597 | (24.7) | 185453 | (24.0) | 1449 | (20.8) |
|  | More than 1 body system | 2019 | (31.9) | 272 | (38.2) | 106714 | (23.0) | 168296 | (21.8) | 1660 | (23.8) |
| Life limiting or non-life limiting comorbidity | Comorbidity present | 2272 | (35.8) | 307 | (43.1) | 169415 | (36.5) | 272959 | (35.4) | 2065 | (29.6) |
|  | Life limiting comorbidity | 1143 | (18.0) | 147 | (20.6) | 51896 | (11.2) | 80790 | (10.5) | 1044 | (15.0) |
| Life limiting neurodisability | Comorbidity present | 170 | (2.7) | <5 | . | 8395 | (1.8) | 13280 | (1.7) | 143 | (2.1) |
| Respiratory comorbidity | **Any respiratory** | 1178 | (18.6) | 105 | (14.7) | 70772 | (15.3) | 131350 | (17.0) | 1328 | (19.1) |
|  | Congenital anomalies | 165 | (2.6) | 8 | (1.1) | 10080 | (2.2) | 17645 | (2.3) | 193 | (2.8) |
|  | Asthma / chronic respiratory | 651 | (10.3) | 49 | (6.9) | 41973 | (9.1) | 79871 | (10.4) | 711 | (10.2) |
|  | Other respiratory | 548 | (8.6) | 64 | (9.0) | 27861 | (6.0) | 52761 | (6.8) | 675 | (9.7) |
|  | Cystic fibrosis | 17 | (0.3) | . | (.) | 1232 | (0.3) | 2348 | (0.3) | 30 | (0.4) |
|  | Injuries (respiratory) | 3 | (0.0) | . | (.) | 111 | (0.0) | 231 | (0.0) | . | (.) |
| Cardiovascular comorbidity | **Any cardiovascular** | 1038 | (16.4) | 286 | (40.2) | 47113 | (10.2) | 72038 | (9.3) | 831 | (11.9) |
|  | Other (cardiac) | 839 | (13.2) | 277 | (38.9) | 34565 | (7.5) | 51464 | (6.7) | 612 | (8.8) |
|  | Congenital | 403 | (6.4) | 38 | (5.3) | 22967 | (5.0) | 37758 | (4.9) | 422 | (6.1) |
| Neurological comorbidity | **Any neurological** | 1248 | (19.7) | 95 | (13.3) | 73785 | (15.9) | 116172 | (15.1) | 1096 | (15.7) |
|  | Injuries (neurological) | 35 | (0.6) | <5 | . | 1714 | (0.4) | 2658 | (0.3) | 25 | (0.4) |
|  | Chronic eye conditions | 206 | (3.3) | 5 | (0.7) | 10776 | (2.3) | 17769 | (2.3) | 190 | (2.7) |
|  | Other (neurological) | 627 | (9.9) | 71 | (10.0) | 34206 | (7.4) | 51757 | (6.7) | 492 | (7.1) |
|  | Cerebral palsy | 179 | (2.8) | 8 | (1.1) | 9018 | (1.9) | 14484 | (1.9) | 181 | (2.6) |
|  | Epilepsy | 557 | (8.8) | 19 | (2.7) | 31959 | (6.9) | 48528 | (6.3) | 451 | (6.5) |
|  | Perinatal conditions | 108 | (1.7) | <5 | . | 7987 | (1.7) | 12514 | (1.6) | 121 | (1.7) |
|  | Chronic ear conditions | 166 | (2.6) | 7 | (1.0) | 9708 | (2.1) | 17451 | (2.3) | 180 | (2.6) |
|  | Congenital (neurological) | 388 | (6.1) | 9 | (1.3) | 19414 | (4.2) | 32187 | (4.2) | 373 | (5.4) |
| Cancer/Haem | **Any Cancer/Haem** | 1007 | (15.9) | 185 | (26.0) | 43933 | (9.5) | 66666 | (8.6) | 795 | (11.4) |
|  | Neoplasms | 351 | (5.5) | 7 | (1.0) | 15523 | (3.3) | 19849 | (2.6) | 260 | (3.7) |
|  | Anaemia / other blood disorders | 930 | (14.7) | 179 | (25.1) | 38973 | (8.4) | 59562 | (7.7) | 713 | (10.2) |
|  | Immunological disorders | 55 | (0.9) | <5 | . | 2311 | (0.5) | 4204 | (0.5) | 73 | (1.0) |
| Met/Endo/Renal/GU Comorbidity | **Any Met/Endo/Renal/GU** | 1446 | (22.8) | 150 | (21.1) | 87235 | (18.8) | 134204 | (17.4) | 1187 | (17.0) |
|  | Digestive | 732 | (11.5) | 44 | (6.2) | 45376 | (9.8) | 73677 | (9.5) | 664 | (9.5) |
|  | Metabolic | 355 | (5.6) | 60 | (8.4) | 14056 | (3.0) | 21637 | (2.8) | 252 | (3.6) |
|  | Diabetes | 154 | (2.4) | <5 | . | 8426 | (1.8) | 11022 | (1.4) | 51 | (0.7) |
|  | Injuries (met/end/renal/GU) | 9 | (0.1) | <5 | . | 343 | (0.1) | 573 | (0.1) | <5 | . |
|  | Renal/GU | 432 | (6.8) | 35 | (4.9) | 25089 | (5.4) | 37730 | (4.9) | 362 | (5.2) |
|  | Other (met/end/renal/GU) | 248 | (3.9) | 28 | (3.9) | 10227 | (2.2) | 14760 | (1.9) | 104 | (1.5) |
|  | Other endocrine | 45 | (0.7) | <5 | . | 2717 | (0.6) | 4676 | (0.6) | 59 | (0.8) |
|  | GI/GU congenital anomalies | 206 | (3.3) | 16 | (2.2) | 15755 | (3.4) | 24645 | (3.2) | 244 | (3.5) |
| MSK/skin Comorbidity | **Any MSK / skin** | 718 | (11.3) | 95 | (13.3) | 37183 | (8.0) | 59605 | (7.7) | 646 | (9.3) |
|  | Congenital anomalies | 173 | (2.7) | 9 | (1.3) | 9015 | (1.9) | 15236 | (2.0) | 191 | (2.7) |
|  | MSK/connective tissue | 551 | (8.7) | 90 | (12.6) | 26139 | (5.6) | 41563 | (5.4) | 431 | (6.2) |
|  | Chronic skin disorders | 128 | (2.0) | <5 | . | 6980 | (1.5) | 10960 | (1.4) | 127 | (1.8) |
|  | Skeletal injuries / amputations | 8 | (0.1) | . | (.) | 1096 | (0.2) | 2044 | (0.3) | 22 | (0.3) |
| Neurological with cardiovascular | | 533 | (8.4) | 67 | (9.4) | 23408 | (5.0) | 37101 | (4.8) | 430 | (6.2) |
| Neurological with respiratory | | 491 | (7.7) | 32 | (4.5) | 24034 | (5.2) | 41928 | (5.4) | 509 | (7.3) |
| Respiratory with cardiovascular | | 434 | (6.8) | 69 | (9.7) | 19351 | (4.2) | 32466 | (4.2) | 434 | (6.2) |

**Table S2** Number and proportion of admissions by selected diagnoses within each cohort (COVID-19, PIMS-TS, other pandemic year admissions; all admissions in 2019/20; influenza admissions 2019/20)

|  | 2020/21 | | | | | | 2019/20 | | | |
| --- | --- | --- | --- | --- | --- | --- | --- | --- | --- | --- |
|  | COVID 19 | | PIMS TS | | Other pandemic year admission | | All admissions 2019/20 | | Influenza 2019/20 | |
|  | n | (%) | n | (%) | n | (%) | n | (%) | n | (%) |
| Epilepsy | 393 | (6.2) | 14 | (2.0) | 20027 | (4.3) | 30502 | (4.0) | 318 | (4.6) |
| Asthma | 618 | (9.8) | 49 | (6.9) | 40740 | (8.8) | 77358 | (10.0) | 681 | (9.8) |
| Sickle Cell Disease | 96 | (1.5) | 8 | (1.1) | 3080 | (0.7) | 5287 | (0.7) | 69 | (1.0) |
| Trisomy 21 | 67 | (1.1) | <5 | . | 2353 | (0.5) | 4437 | (0.6) | 49 | (0.7) |
| Diabetes Mellitus | 154 | (2.4) | <5 | . | 8382 | (1.8) | 10935 | (1.4) | 50 | (0.7) |

Table S3 Total admissions and proportion resulting in paediatric intensive care admission (PICU) by sociodemographic characteristics within each cohort (COVID-19, PIMS-TS, other non-traumatic admissions in 2020/21; all non-traumatic admissions and those due to influenza in 2019/20)

|  |  | 2020/21 | | | | | | | | | 2019/20 | | | | | |
| --- | --- | --- | --- | --- | --- | --- | --- | --- | --- | --- | --- | --- | --- | --- | --- | --- |
|  |  | COVID 19 | | | PIMS TS | | | Other pandemic year admission | | | All admissions 2019/20 | | | Influenza 2019/20 | | |
|  |  | n | PICU | % PICU | n | PICU | % PICU | n | PICU | % PICU | n | PICU | % PICU | n | PICU | % PICU |
|  | Total | 6338 | 259 | (4.1) | 712 | 312 | (43.8) | 463556 | 5016 | (1.1) | 771591 | 7282 | (0.9) | 6968 | 161 | (2.3) |
| Sex | Male | 3347 | 149 | (4.5) | 452 | 181 | (40.0) | 247299 | 2830 | (1.1) | 416830 | 4212 | (1.0) | 3733 | 94 | (2.5) |
|  | Female | 2991 | 110 | (3.7) | 260 | 131 | (50.4) | 216257 | 2186 | (1.0) | 354761 | 3071 | (0.9) | 3235 | 67 | (2.1) |
| Age | Neonates | 741 | 29 | (3.9) | <5 | 0 | (0.0) | 69230 | 1280 | (1.8) | 89822 | 1618 | (1.8) | 151 | 8 | (5.3) |
|  | Post neonatal | 1216 | 35 | (2.9) | 71 | 10 | (14.1) | 71560 | 928 | (1.3) | 135195 | 1702 | (1.3) | 1036 | 25 | (2.4) |
|  | 1 to 4 | 1281 | 50 | (3.9) | 217 | 47 | (21.7) | 126426 | 1198 | (0.9) | 262511 | 1946 | (0.7) | 3189 | 65 | (2.0) |
|  | 5 to 9 | 840 | 46 | (5.5) | 216 | 111 | (51.4) | 71255 | 692 | (1.0) | 116951 | 956 | (0.8) | 1274 | 32 | (2.5) |
|  | 10 to 14 | 1188 | 64 | (5.4) | 175 | 120 | (68.6) | 72256 | 630 | (0.9) | 97662 | 806 | (0.8) | 871 | 24 | (2.8) |
|  | 15 to 17 | 1072 | 35 | (3.3) | 31 | 24 | (77.4) | 52829 | 288 | (0.5) | 69450 | 255 | (0.4) | 447 | 7 | (1.6) |
| Ethnicity | White | 3685 | 141 | (3.8) | 285 | 93 | (32.6) | 329358 | 3054 | (0.9) | 544460 | 4644 | (0.9) | 4653 | 101 | (2.2) |
|  | Mixed | 279 | 5 | (1.8) | 51 | 20 | (39.2) | 20426 | 214 | (1.0) | 33397 | 325 | (1.0) | 259 | 6 | (2.3) |
|  | Asian | 1203 | 50 | (4.2) | 155 | 79 | (51.0) | 49217 | 736 | (1.5) | 88615 | 1006 | (1.1) | 1063 | 25 | (2.4) |
|  | Black | 395 | 33 | (8.4) | 109 | 62 | (56.9) | 17388 | 353 | (2.0) | 30948 | 514 | (1.7) | 333 | 10 | (3.0) |
|  | Other | 298 | 12 | (4.0) | 40 | 19 | (47.5) | 13160 | 164 | (1.2) | 22381 | 260 | (1.2) | 197 | 8 | (4.1) |
|  | Unknown | 478 | 18 | (3.8) | 72 | 39 | (54.2) | 34007 | 495 | (1.5) | 51790 | 534 | (1.0) | 463 | 11 | (2.4) |
| IMD Category | Most deprived | 1662 | 71 | (4.3) | 163 | 61 | (37.4) | 108096 | 1294 | (1.2) | 188391 | 1946 | (1.0) | 1906 | 50 | (2.6) |
|  | 2nd most deprived | 1533 | 71 | (4.6) | 182 | 88 | (48.4) | 96386 | 1158 | (1.2) | 163405 | 1596 | (1.0) | 1497 | 33 | (2.2) |
|  | 3rd most deprived | 1218 | 45 | (3.7) | 164 | 82 | (50.0) | 92034 | 942 | (1.0) | 152297 | 1403 | (0.9) | 1278 | 30 | (2.3) |
|  | 4th most deprived | 1087 | 35 | (3.2) | 98 | 38 | (38.8) | 87873 | 852 | (1.0) | 142097 | 1259 | (0.9) | 1197 | 27 | (2.3) |
|  | Least deprived | 838 | 37 | (4.4) | 105 | 43 | (41.0) | 78982 | 768 | (1.0) | 125167 | 1074 | (0.9) | 1090 | 21 | (1.9) |
|  | Missing | . | . | (.) | . | . | (.) | 185 | <5 | . | 234 | 5 | (2.1) | . | . | (.) |

| **Table S4** Total admissions and proportion resulting in paediatric intensive care admission (PICU) by selected body system comorbidity within each cohort (COVID-19, PIMS-TS, other non-traumatic admissions in 2020/21; all non-traumatic admissions and those due to influenza in 2019/20) | | | | | | | | | | | | | | | | |
| --- | --- | --- | --- | --- | --- | --- | --- | --- | --- | --- | --- | --- | --- | --- | --- | --- |
|  |  | COVID 19 | | | PIMS TS | | | Other pandemic year admission | | | All admissions 2019/20 | | | Influenza 2019/20 | | |
|  |  | n | PICU | % PICU | n | PICU | % PICU | n | PICU | % PICU | n | PICU | % PICU | n | PICU | % PICU |
| Any comorbidity | No comorbidity | 2923 | 22 | (0.8) | 258 | 50 | (19.4) | 242245 | 358 | (0.1) | 417842 | 811 | (0.2) | 3859 | 30 | (0.8) |
|  | Comorbidity present | 3415 | 237 | (6.9) | 454 | 262 | (57.7) | 221311 | 4658 | (2.1) | 353749 | 6472 | (1.8) | 3109 | 131 | (4.2) |
| Number of body systems | 1 body system | 1396 | 35 | (2.5) | 182 | 76 | (41.8) | 114597 | 1015 | (0.9) | 185453 | 1436 | (0.8) | 1449 | 29 | (2.0) |
|  | More than 1 body system | 2019 | 202 | (10.0) | 272 | 186 | (68.4) | 106714 | 3643 | (3.4) | 168296 | 5036 | (3.0) | 1660 | 102 | (6.1) |
| Life limiting or non-life limiting comorbidity | Comorbidity present | 2272 | 79 | (3.5) | 307 | 157 | (51.1) | 169415 | 1701 | (1.0) | 272959 | 2323 | (0.9) | 2065 | 43 | (2.1) |
|  | Life limiting comorbidity | 1143 | 158 | (13.8) | 147 | 105 | (71.4) | 51896 | 2957 | (5.7) | 80790 | 4149 | (5.1) | 1044 | 88 | (8.4) |
| Life limiting neurodisability | Comorbidity present | 170 | 28 | (16.5) | <5 | <5 | (100.0) | 8395 | 473 | (5.6) | 13280 | 718 | (5.4) | 143 | 20 | (14.0) |
| Respiratory comorbidity | **Any respiratory** | 1178 | 122 | (10.4) | 105 | 66 | (62.9) | 70772 | 2217 | (3.1) | 131350 | 3607 | (2.7) | 1328 | 95 | (7.2) |
|  | Congenital anomalies | 165 | 17 | (10.3) | 8 | 5 | (62.5) | 10080 | 581 | (5.8) | 17645 | 883 | (5.0) | 193 | 18 | (9.3) |
|  | Asthma and chronic respiratory conditions | 651 | 34 | (5.2) | 49 | 26 | (53.1) | 41973 | 408 | (1.0) | 79871 | 735 | (0.9) | 711 | 17 | (2.4) |
|  | Other respiratory | 548 | 97 | (17.7) | 64 | 49 | (76.6) | 27861 | 1802 | (6.5) | 52761 | 3063 | (5.8) | 675 | 85 | (12.6) |
|  | Cystic fibrosis | 17 | <5 | . | . | . | (.) | 1232 | 23 | (1.9) | 2348 | 30 | (1.3) | 30 | 0 | (0.0) |
|  | Injuries (respiratory) | 3 | <5 | . | . | . | (.) | 111 | 9 | (8.1) | 231 | 16 | (6.9) | . | . | (.) |
| Cardiovascular comorbidity | **Any cardiovascular** | 1038 | 155 | (14.9) | 286 | 180 | (62.9) | 47113 | 2855 | (6.1) | 72038 | 3718 | (5.2) | 831 | 56 | (6.7) |
|  | Other (cardiac) | 839 | 133 | (15.9) | 277 | 175 | (63.2) | 34565 | 2261 | (6.5) | 51464 | 3006 | (5.8) | 612 | 42 | (6.9) |
|  | Congenital heart disease | 403 | 60 | (14.9) | 38 | 19 | (50.0) | 22967 | 1687 | (7.3) | 37758 | 2265 | (6.0) | 422 | 33 | (7.8) |
| Neurological comorbidity | **Any neurological** | 1248 | 149 | (11.9) | 95 | 62 | (65.3) | 73785 | 2651 | (3.6) | 116172 | 3762 | (3.2) | 1096 | 85 | (7.8) |
|  | Injuries (neurological) | 35 | 5 | (14.3) | <5 | . | . | 1714 | 56 | (3.3) | 2658 | 82 | (3.1) | 25 | 0 | (0.0) |
|  | Chronic eye conditions | 206 | 32 | (15.5) | <5 | <5 | . | 10776 | 566 | (5.3) | 17769 | 806 | (4.5) | 190 | 22 | (11.6) |
|  | Other (neurological) | 627 | 88 | (14.0) | 71 | 51 | (71.8) | 34206 | 1572 | (4.6) | 51757 | 2215 | (4.3) | 492 | 50 | (10.2) |
|  | Cerebral palsy | 179 | 33 | (18.4) | 8 | 6 | (75.0) | 9018 | 493 | (5.5) | 14484 | 767 | (5.3) | 181 | 22 | (12.2) |
|  | Epilepsy | 557 | 76 | (13.6) | 19 | 12 | (63.2) | 31959 | 1234 | (3.9) | 48528 | 1814 | (3.7) | 451 | 52 | (11.5) |
|  | Perinatal conditions | 108 | 11 | (10.2) | <5 | <5 | (100.0) | 7987 | 529 | (6.6) | 12514 | 683 | (5.5) | 121 | 13 | (10.7) |
|  | Chronic ear conditions | 166 | 20 | (12.0) | 7 | <5 | . | 9708 | 298 | (3.1) | 17451 | 411 | (2.4) | 180 | 9 | (5.0) |
|  | Congenital (neurological) | 388 | 58 | (14.9) | 9 | 5 | (55.6) | 19414 | 908 | (4.7) | 32187 | 1360 | (4.2) | 373 | 32 | (8.6) |
| Cancer/Haema | **Any Cancer/Haem** | 1007 | 95 | (9.4) | 185 | 133 | (71.9) | 43933 | 1514 | (3.4) | 66666 | 2039 | (3.1) | 795 | 40 | (5.0) |
|  | Neoplasms | 351 | 33 | (9.4) | 7 | <5 | . | 15523 | 509 | (3.3) | 19849 | 564 | (2.8) | 260 | <5 | . |
|  | Anaemia and other blood disorders | 930 | 91 | (9.8) | 179 | 131 | (73.2) | 38973 | 1306 | (3.4) | 59562 | 1837 | (3.1) | 713 | 39 | (5.5) |
|  | Immunological disorders | 55 | <5 | . | <5 | <5 | . | 2311 | 100 | (4.3) | 4204 | 143 | (3.4) | 73 | 6 | (8.2) |
| Met/Endo/Renal/GU Comorbidity | **Any Met/Endo/Renal/GU** | 1446 | 114 | (7.9) | 150 | 99 | (66.0) | 87235 | 2288 | (2.6) | 134204 | 3187 | (2.4) | 1187 | 63 | (5.3) |
|  | Digestive | 732 | 48 | (6.6) | 44 | 30 | (68.2) | 45376 | 1153 | (2.5) | 73677 | 1750 | (2.4) | 664 | 24 | (3.6) |
|  | Metabolic | 355 | 44 | (12.4) | 60 | 41 | (68.3) | 14056 | 702 | (5.0) | 21637 | 1039 | (4.8) | 252 | 19 | (7.5) |
|  | Diabetes | 154 | 16 | (10.4) | <5 | . | . | 8426 | 207 | (2.5) | 11022 | 172 | (1.6) | 51 | <5 | . |
|  | Injuries (met/end/renal/GU) | 9 | 0 | (0.0) | <5 | 0 | (0.0) | 343 | 22 | (6.4) | 573 | 25 | (4.4) | <5 | 0 | (0.0) |
|  | Renal/GU | 432 | 46 | (10.6) | 35 | 27 | (77.1) | 25089 | 839 | (3.3) | 37730 | 1190 | (3.2) | 362 | 20 | (5.5) |
|  | Other (met/end/renal/GU) | 248 | 26 | (10.5) | 28 | 24 | (85.7) | 10227 | 247 | (2.4) | 14760 | 330 | (2.2) | 104 | 6 | (5.8) |
|  | Other endocrine | 45 | 5 | (11.1) | <5 | . | (100.0) | 2717 | 108 | (4.0) | 4676 | 140 | (3.0) | 59 | 7 | (11.9) |
|  | Congenital anomalies of GI/GU system | 206 | 18 | (8.7) | 16 | 9 | (56.3) | 15755 | 624 | (4.0) | 24645 | 896 | (3.6) | 244 | 14 | (5.7) |
| MSK/skin Comorbidity | **Any MSK/skin** | 718 | 74 | (10.3) | 95 | 62 | (65.3) | 37183 | 1063 | (2.9) | 59605 | 1635 | (2.7) | 646 | 39 | (6.0) |
|  | Congenital anomalies | 173 | 22 | (12.7) | 9 | 5 | (55.6) | 9015 | 408 | (4.5) | 15236 | 582 | (3.8) | 191 | 15 | (7.9) |
|  | MSK/connective tissue | 551 | 54 | (9.8) | 90 | 61 | (67.8) | 26139 | 691 | (2.6) | 41563 | 1150 | (2.8) | 431 | 30 | (7.0) |
|  | Chronic skin disorders | 128 | 15 | (11.7) | <5 | <5 | . | 6980 | 269 | (3.9) | 10960 | 350 | (3.2) | 127 | 5 | (3.9) |
|  | Skeletal injuries / amputations | 8 | 0 | (0.0) | . | . | (.) | 1096 | 25 | (2.3) | 2044 | 55 | (2.7) | 22 | <5 | . |
| Neurological and cardiovascular | | 533 | 98 | (18.4) | 67 | 50 | (74.6) | 23408 | 1645 | (7.0) | 37101 | 2288 | (6.2) | 430 | 38 | (8.8) |
| Neurological and respiratory | | 491 | 86 | (17.5) | 32 | 25 | (78.1) | 24034 | 1370 | (5.7) | 41928 | 2254 | (5.4) | 509 | 60 | (11.8) |
| Respiratory and cardiovascular | | 434 | 85 | (19.6) | 69 | 50 | (72.5) | 19351 | 1431 | (7.4) | 32466 | 2169 | (6.7) | 434 | 45 | (10.4) |

**Table S5** Total admissions and proportion resulting in paediatric intensive care admission (PICU) by selected diagnoses within each cohort (COVID-19, PIMS-TS, other non-traumatic admissions in 2020/21; all non-traumatic admissions and those due to influenza in 2019/20)

|  | 2020/21 | | | | | | | | | 2019/20 | | | | | |
| --- | --- | --- | --- | --- | --- | --- | --- | --- | --- | --- | --- | --- | --- | --- | --- |
|  | COVID 19 | | | PIMS TS | | | Other pandemic year admission | | | All admissions 2019/20 | | | Influenza 2019/20 | | |
|  | n | PICU | % PICU | n | PICU | % PICU | n | PICU | % PICU | n | PICU | % PICU | n | PICU | % PICU |
| Epilepsy | 618 | 31 | (5.0) | 49 | 26 | (53.1) | 40740 | 359 | (0.9) | 77358 | 603 | (0.8) | 681 | 16 | (2.3) |
| Asthma | 96 | <5 | . | 8 | 7 | (87.5) | 3080 | 53 | (1.7) | 5287 | 91 | (1.7) | 69 | <5 | . |
| Sickle Cell Disease | 67 | 16 | (23.9) | <5 | 0 | (0.0) | 2353 | 112 | (4.8) | 4437 | 214 | (4.8) | 49 | <5 | . |
| Trisomy 21 | 154 | 16 | (10.4) | <5 | <5 | . | 8382 | 205 | (2.4) | 10935 | 171 | (1.6) | 50 | <5 | . |
| Diabetes Mellitus | 618 | 31 | (5.0) | 49 | 26 | (53.1) | 40740 | 359 | (0.9) | 77358 | 603 | (0.8) | 681 | 16 | (2.3) |

Table S6 Output from univariable analyses: Odds ratios (OR) with 95% confidence intervals (CI) of admission to PICU by sociodemographic characteristics within the COVID-19 cohort

|  |  | OR | lower CI | upper CI | p |
| --- | --- | --- | --- | --- | --- |
| Sex | Male (baseline) | 1 |  |  |  |
|  | Female | 0.82 | 0.63 | 1.06 | 0.125 |
| Age | 1 to 4 (baseline) |  |  |  |  |
|  | Neonates | 1.04 | 0.64 | 1.70 | 0.874 |
|  | Post neonatal Infants | 0.78 | 0.49 | 1.23 | 0.288 |
|  | 5 to 9 | 1.50 | 0.98 | 2.30 | 0.063 |
|  | 10 to 14 | 1.50 | 1.01 | 2.22 | 0.045 |
|  | 15 to 17 | 0.89 | 0.57 | 1.40 | 0.620 |
| Ethnicity | White | 1 |  |  |  |
|  | Mixed | 0.48 | 0.19 | 1.18 | 0.110 |
|  | Asian | 1.10 | 0.79 | 1.54 | 0.571 |
|  | Black | 2.35 | 1.58 | 3.49 | 0.000 |
|  | Other | 1.10 | 0.60 | 2.01 | 0.760 |
|  | Unknown | 0.95 | 0.55 | 1.62 | 0.843 |
| IMD Category | Most deprived | 1 |  |  |  |
|  | 2nd most deprived | 1.07 | 0.76 | 1.50 | 0.715 |
|  | 3rd most deprived | 0.86 | 0.59 | 1.26 | 0.432 |
|  | 4th most deprived | 0.72 | 0.47 | 1.10 | 0.127 |
|  | Least deprived | 1.00 | 0.66 | 1.52 | 0.999 |

**Table S7** Output from univariable analyses: Odds ratios (OR) with 95% confidence intervals (CI) of admission to PICU by sociodemographic characteristics within the PIMS-TS cohort

|  |  | OR | lower CI | upper CI | p |
| --- | --- | --- | --- | --- | --- |
| Sex | Male (baseline) |  |  |  |  |
|  | Female | 1.48 | 1.08 | 2.02 | 0.014 |
| Age | 1 to 4 (baseline) | 1 |  |  |  |
|  | Neonates | - | - | - | - |
|  | Post neonatal Infants | 0.64 | 0.30 | 1.35 | 0.237 |
|  | 5 to 9 | 3.98 | 2.60 | 6.10 | 0.000 |
|  | 10 to 14 | 8.30 | 5.23 | 13.18 | 0.000 |
|  | 15 to 17 | 12.61 | 5.10 | 31.17 | 0.000 |
| Ethnicity | White | 1 |  |  |  |
|  | Mixed | 1.42 | 0.76 | 2.66 | 0.273 |
|  | Asian | 2.16 | 1.45 | 3.24 | 0.000 |
|  | Black | 2.74 | 1.73 | 4.32 | 0.000 |
|  | Other | 1.91 | 0.98 | 3.74 | 0.059 |
|  | Unknown | 2.43 | 1.42 | 4.17 | 0.001 |
| IMD Category | Most deprived | 1 |  |  |  |
|  | 2nd most deprived | 1.53 | 0.99 | 2.36 | 0.058 |
|  | 3rd most deprived | 1.61 | 1.03 | 2.51 | 0.037 |
|  | 4th most deprived | 1.02 | 0.61 | 1.70 | 0.954 |
|  | Least deprived | 1.10 | 0.66 | 1.83 | 0.710 |

**Table S8** Output from univariable analyses: Odds ratios (OR) with 95% confidence intervals (CI) of admission to PICU by sociodemographic characteristics within the Other pandemic year admissions cohort

|  |  | OR | lower CI | upper CI | p |
| --- | --- | --- | --- | --- | --- |
| Sex | Male (baseline) | 1 |  |  |  |
|  | Female | 0.89 | 0.83 | 0.94 | 0.000 |
| Age | 1 to 4 (baseline) |  |  |  |  |
|  | Neonates | 2.27 | 2.09 | 2.47 | 0.00 |
|  | Post neonatal Infants | 1.33 | 1.20 | 1.46 | 0.00 |
|  | 5 to 9 | 1.03 | 0.93 | 1.14 | 0.55 |
|  | 10 to 14 | 0.97 | 0.87 | 1.08 | 0.55 |
|  | 15 to 17 | 0.60 | 0.52 | 0.69 | 0.00 |
| Ethnicity | White | 1 |  |  |  |
|  | Mixed | 1.13 | 0.97 | 1.31 | 0.123 |
|  | Asian | 1.67 | 1.53 | 1.82 | 0.000 |
|  | Black | 2.29 | 2.03 | 2.58 | 0.000 |
|  | Other | 1.41 | 1.20 | 1.66 | 0.000 |
|  | Unknown | 1.66 | 1.50 | 1.84 | 0.000 |
| IMD Category | Most deprived | 1 |  |  |  |
|  | 2nd most deprived | 1.01 | 0.93 | 1.10 | 0.796 |
|  | 3rd most deprived | 0.85 | 0.78 | 0.94 | 0.001 |
|  | 4th most deprived | 0.80 | 0.73 | 0.88 | 0.000 |
|  | Least deprived | 0.81 | 0.74 | 0.89 | 0.000 |

**Table S9** Output from univariable analyses: Odds ratios (OR) with 95% confidence intervals (CI) of admission to PICU by sociodemographic characteristics within the all admissions 2019/20

|  |  | OR | lower CI | upper CI | p |
| --- | --- | --- | --- | --- | --- |
| Sex | Male (baseline) | 1 |  |  |  |
|  | Female | 0.84 | 0.80 | 0.89 | 0.000 |
| Age | 1 to 4 (baseline) | 1 |  |  |  |
|  | Neonates | 2.89 | 2.69 | 3.11 | 0.000 |
|  | Post neonatal Infants | 1.71 | 1.58 | 1.85 | 0.000 |
|  | 5 to 9 | 1.11 | 1.02 | 1.22 | 0.016 |
|  | 10 to 14 | 1.17 | 1.07 | 1.28 | 0.001 |
|  | 15 to 17 | 0.54 | 0.46 | 0.62 | 0.000 |
| Ethnicity | White | 1 |  |  |  |
|  | Mixed | 1.12 | 0.99 | 1.27 | 0.065 |
|  | Asian | 1.33 | 1.23 | 1.44 | 0.000 |
|  | Black | 2.02 | 1.83 | 2.24 | 0.000 |
|  | Other | 1.33 | 1.16 | 1.52 | 0.000 |
|  | Unknown | 1.25 | 1.14 | 1.37 | 0.000 |
| IMD Category | Most deprived | 1 |  |  |  |
|  | 2nd most deprived | 0.96 | 0.89 | 1.03 | 0.212 |
|  | 3rd most deprived | 0.90 | 0.83 | 0.97 | 0.005 |
|  | 4th most deprived | 0.86 | 0.79 | 0.93 | 0.000 |
|  | Least deprived | 0.84 | 0.78 | 0.92 | 0.000 |

**Table S10** Output from univariable analyses: Odds ratios (OR) with 95% confidence intervals (CI) of admission to PICU by sociodemographic characteristics within Influenza admissions 2019/20

|  |  | OR | lower CI | upper CI | p |
| --- | --- | --- | --- | --- | --- |
| Sex | Male (baseline) | 1 |  |  |  |
|  | Female | 0.82 | 0.59 | 1.12 | 0.211 |
| Age | 1 to 4 (baseline) | 1 |  |  |  |
|  | Neonates | 2.71 | 1.28 | 5.76 | 0.009 |
|  | Post neonatal Infants | 1.20 | 0.75 | 1.92 | 0.444 |
|  | 5 to 9 | 1.25 | 0.81 | 1.92 | 0.306 |
|  | 10 to 14 | 1.37 | 0.85 | 2.20 | 0.196 |
|  | 15 to 17 | 0.77 | 0.35 | 1.68 | 0.508 |
| Ethnicity | White | 1 |  |  |  |
|  | Mixed | 1.07 | 0.47 | 2.47 | 0.871 |
|  | Asian | 1.10 | 0.70 | 1.71 | 0.685 |
|  | Black | 1.40 | 0.72 | 2.70 | 0.321 |
|  | Other | 1.91 | 0.92 | 3.98 | 0.085 |
|  | Unknown | 1.10 | 0.59 | 2.07 | 0.765 |
| IMD Category | Most deprived | 1 |  |  |  |
|  | 2nd most deprived | 0.84 | 0.54 | 1.31 | 0.438 |
|  | 3rd most deprived | 0.90 | 0.57 | 1.42 | 0.634 |
|  | 4th most deprived | 0.85 | 0.53 | 1.38 | 0.509 |
|  | Least deprived | 0.73 | 0.44 | 1.22 | 0.229 |

**Table S11** Output from multivariable analyses: Odds ratios (OR) with 95% confidence intervals (CI) of admission to PICU by sociodemographic characteristics and co-morbidities by body system number within COVID-19 admissions (full model)

|  |  | OR | lower CI | upper CI | p |
| --- | --- | --- | --- | --- | --- |
| Observations | 6338 |  |  |  |  |
| Sex | Male (baseline) | 1 |  |  |  |
|  | Female | 0.87 | 0.67 | 1.14 | 0.316 |
| Age | 1 to 4 (baseline) | 1 |  |  |  |
|  | Neonates | 2.70 | 1.56 | 4.68 | 0.000 |
|  | Post neonatal Infants | 1.32 | 0.82 | 2.14 | 0.259 |
|  | 5 to 9 | 1.09 | 0.70 | 1.71 | 0.690 |
|  | 10 to 14 | 1.15 | 0.77 | 1.73 | 0.502 |
|  | 15 to 17 | 0.58 | 0.36 | 0.92 | 0.021 |
| Ethnicity | White | 1 |  |  |  |
|  | Mixed | 0.54 | 0.22 | 1.35 | 0.185 |
|  | Asian | 0.93 | 0.65 | 1.32 | 0.677 |
|  | Black | 1.89 | 1.23 | 2.90 | 0.004 |
|  | Other | 0.94 | 0.50 | 1.76 | 0.846 |
|  | Unknown | 0.99 | 0.57 | 1.72 | 0.974 |
| IMD Category | Most deprived | 1 |  |  |  |
|  | 2nd most deprived | 1.13 | 0.80 | 1.61 | 0.485 |
|  | 3rd most deprived | 1.02 | 0.68 | 1.51 | 0.933 |
|  | 4th most deprived | 0.91 | 0.58 | 1.42 | 0.668 |
|  | Least deprived | 1.33 | 0.85 | 2.10 | 0.212 |
| Number of comorbidities | No comorbidities | 1 |  |  |  |
|  | 1 body system | 4.14 | 2.34 | 7.34 | 0.000 |
|  | More than 1 body system | 20.07 | 12.13 | 33.22 | 0.000 |
| Constant |  | 0.01 | 0.00 | 0.01 | 0.000 |

**Table S12** Output from multivariable analyses: Odds ratios (OR) with 95% confidence intervals (CI) of admission to PICU by sociodemographic characteristics and co-morbidities by body system number within PIMS-TS admissions (full model)

|  |  | OR | lower CI | upper CI | p |
| --- | --- | --- | --- | --- | --- |
| Observations | 710 |  |  |  |  |
| Sex | Male (baseline) | 1 |  |  |  |
|  | Female | 1.75 | 1.19 | 2.56 | 0.004 |
| Age | 1 to 4 (baseline) | 1 |  |  |  |
|  | Neonates | - | - | - |  |
|  | Post neonatal Infants | 0.50 | 0.23 | 1.11 | 0.088 |
|  | 5 to 9 | 3.12 | 1.96 | 4.98 | 0.000 |
|  | 10 to 14 | 5.74 | 3.47 | 9.49 | 0.000 |
|  | 15 to 17 | 6.89 | 2.82 | 16.83 | 0.000 |
| Ethnicity | White (baseline) | 1 |  |  |  |
|  | Mixed | 1.49 | 0.69 | 3.20 | 0.312 |
|  | Asian | 2.03 | 1.25 | 3.28 | 0.004 |
|  | Black | 2.20 | 1.21 | 4.01 | 0.010 |
|  | Other | 1.88 | 0.79 | 4.46 | 0.151 |
|  | Unknown | 2.18 | 1.16 | 4.13 | 0.016 |
| IMD Category | Most deprived (baseline) | 1 |  |  |  |
|  | 2nd most deprived | 1.69 | 0.98 | 2.92 | 0.060 |
|  | 3rd most deprived | 1.87 | 1.09 | 3.23 | 0.024 |
|  | 4th most deprived | 1.18 | 0.64 | 2.19 | 0.590 |
|  | Least deprived | 1.47 | 0.75 | 2.90 | 0.262 |
| Number of comorbidities | No comorbidities (baseline) | 1 |  |  |  |
|  | 1 body system | 2.59 | 1.61 | 4.15 | 0.000 |
|  | More than 1 body system | 6.93 | 4.42 | 10.88 | 0.000 |
| Constant |  | 0.04 | 0.02 | 0.08 | 0.000 |

**Table S13** Output from multivariable analyses: Odds ratios (OR) with 95% confidence intervals (CI) of admission to PICU by sociodemographic characteristics and co-morbidities by body system number within all other pandemic year admissions (full model)

|  |  | OR | lower CI | upper CI | p |
| --- | --- | --- | --- | --- | --- |
| Observations | 463371 |  |  |  |  |
| Sex | Male (baseline) | 1 |  |  |  |
|  | Female | 0.95 | 0.90 | 1.01 | 0.128 |
| Age | 1 to 4 (baseline) |  |  |  |  |
|  | Neonates | 5.05 | 4.62 | 5.52 | 0.000 |
|  | Post neonatal Infants | 1.68 | 1.54 | 1.84 | 0.000 |
|  | 5 to 9 | 0.74 | 0.67 | 0.82 | 0.000 |
|  | 10 to 14 | 0.63 | 0.57 | 0.70 | 0.000 |
|  | 15 to 17 | 0.32 | 0.28 | 0.37 | 0.000 |
| Ethnicity | White (baseline) |  |  |  |  |
|  | Mixed | 1.16 | 0.99 | 1.35 | 0.065 |
|  | Asian | 1.54 | 1.41 | 1.68 | 0.000 |
|  | Black | 2.08 | 1.84 | 2.36 | 0.000 |
|  | Other | 1.51 | 1.28 | 1.78 | 0.000 |
|  | Unknown | 1.57 | 1.42 | 1.74 | 0.000 |
| IMD Category | Most deprived (baseline) |  |  |  |  |
|  | 2nd most deprived | 1.07 | 0.98 | 1.16 | 0.135 |
|  | 3rd most deprived | 0.97 | 0.89 | 1.07 | 0.558 |
|  | 4th most deprived | 1.02 | 0.92 | 1.12 | 0.695 |
|  | Least deprived | 1.11 | 1.01 | 1.23 | 0.031 |
| Number of comorbidities | No comorbidities (baseline) |  |  |  |  |
|  | 1 body system | 9.67 | 8.53 | 10.96 | 0.000 |
|  | More than 1 body system | 48.58 | 43.14 | 54.71 | 0.000 |
| Constant |  | 0.0007 | 0.0006 | 0.0008 | 0.000 |

**Table S14** Output from multivariable analyses: Odds ratios (OR) with 95% confidence intervals (CI) of admission to PICU by sociodemographic characteristics and co-morbidities by body system number within all admissions 2019/20 (full model)

|  |  | OR | lower CI | upper CI | p |
| --- | --- | --- | --- | --- | --- |
| Observations | 771357 |  |  |  |  |
| Sex | Male (baseline) | 1 |  |  |  |
|  | Female | 0.94 | 0.89 | 0.99 | 0.011 |
| Age | 1 to 4 (baseline) | 1 |  |  |  |
|  | Neonates | 5.14 | 4.77 | 5.52 | 0.000 |
|  | Post neonatal Infants | 2.16 | 2.01 | 2.31 | 0.000 |
|  | 5 to 9 | 0.83 | 0.76 | 0.90 | 0.000 |
|  | 10 to 14 | 0.72 | 0.66 | 0.79 | 0.000 |
|  | 15 to 17 | 0.28 | 0.24 | 0.32 | 0.000 |
| Ethnicity | White (baseline) | 1 |  |  |  |
|  | Mixed | 1.15 | 1.02 | 1.30 | 0.028 |
|  | Asian | 1.30 | 1.21 | 1.40 | 0.000 |
|  | Black | 1.91 | 1.72 | 2.11 | 0.000 |
|  | Other | 1.44 | 1.26 | 1.64 | 0.000 |
|  | Unknown | 1.33 | 1.21 | 1.46 | 0.000 |
| IMD Category | Most deprived (baseline) | 1 |  |  |  |
|  | 2nd most deprived | 0.99 | 0.92 | 1.07 | 0.862 |
|  | 3rd most deprived | 1.01 | 0.94 | 1.09 | 0.767 |
|  | 4th most deprived | 1.03 | 0.96 | 1.12 | 0.402 |
|  | Least deprived | 1.08 | 0.99 | 1.17 | 0.076 |
| Number of comorbidities | No comorbidities (baseline) | 1 |  |  |  |
|  | 1 body system | 5.73 | 5.24 | 6.27 | 0.000 |
|  | More than 1 body system | 27.58 | 25.45 | 29.89 | 0.000 |
| Constant |  |  |  |  |  |

**Table S15** Output from multivariable analyses: Odds ratios (OR) with 95% confidence intervals (CI) of admission to PICU by sociodemographic characteristics and co-morbidities by body system number within influenza admissions 2019/20 (full model)

|  |  | OR | lower CI | upper CI | p |
| --- | --- | --- | --- | --- | --- |
| Observations | 6968 |  |  |  |  |
| Sex | Male (baseline) |  |  |  |  |
|  | Female | 0.94 | 0.68 | 1.30 | 0.692 |
| Age | 1 to 4 (baseline) | 1 |  |  |  |
|  | Neonates | 3.52 | 1.56 | 7.96 | 0.003 |
|  | Post neonatal Infants | 1.40 | 0.87 | 2.25 | 0.164 |
|  | 5 to 9 | 0.90 | 0.58 | 1.40 | 0.634 |
|  | 10 to 14 | 1.02 | 0.63 | 1.64 | 0.946 |
|  | 15 to 17 | 0.58 | 0.26 | 1.27 | 0.173 |
| Ethnicity | White (baseline) | 1 |  |  |  |
|  | Mixed | 1.05 | 0.45 | 2.44 | 0.912 |
|  | Asian | 0.97 | 0.62 | 1.52 | 0.887 |
|  | Black | 1.19 | 0.60 | 2.36 | 0.626 |
|  | Other | 1.70 | 0.81 | 3.58 | 0.164 |
|  | Unknown | 1.20 | 0.62 | 2.29 | 0.591 |
| IMD Category | Most deprived (baseline) |  |  |  |  |
|  | 2nd most deprived | 0.88 | 0.55 | 1.39 | 0.577 |
|  | 3rd most deprived | 0.96 | 0.61 | 1.52 | 0.860 |
|  | 4th most deprived | 0.91 | 0.56 | 1.48 | 0.702 |
|  | Least deprived | 0.92 | 0.54 | 1.56 | 0.756 |
| Number of comorbidities | No comorbidities (baseline) | 1 |  |  |  |
|  | 1 body system | 2.82 | 1.68 | 4.72 | 0.000 |
|  | More than 1 body system | 9.18 | 6.06 | 13.92 | 0.000 |
| Constant |  | 0.01 | 0.00 | 0.01 | 0.000 |

Figure S1 Odds ratios and percentage point difference in predicted probability with 95% confidence intervals for admission to PICU by comorbidity groups within each cohort, adjusted for sex, IMD category, ethnicity in 11-17 year olds only

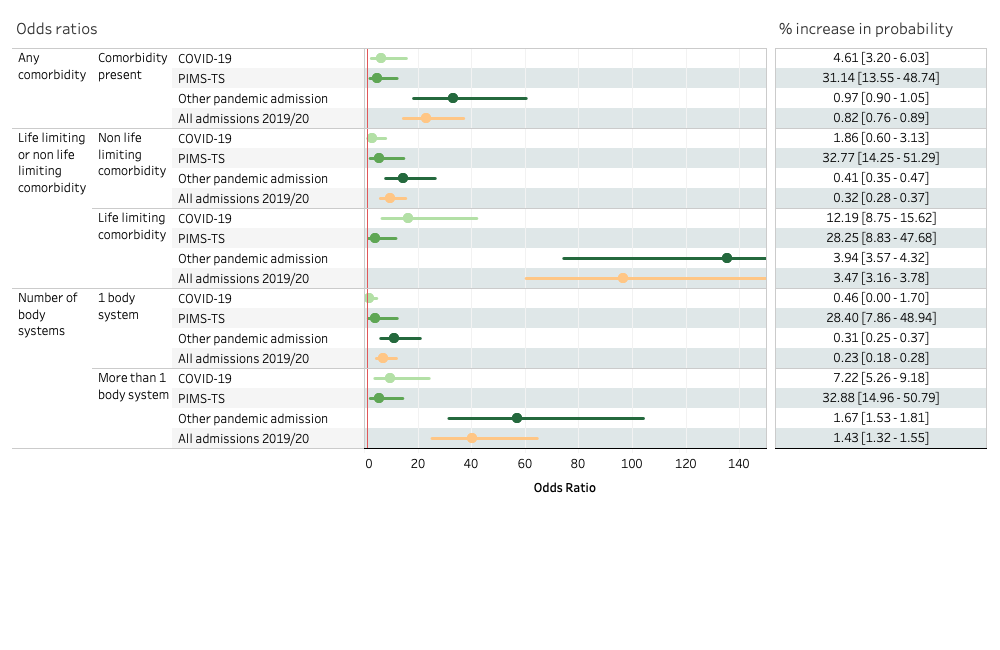

Notes: Results compare odds of PICU admission in selected comorbidity compared with no comorbidity. Upper limits for confidence intervals have been truncated for selected comorbidities for clarity. The red line indicates odds ratio of 1. Percentage point difference in predicted probability of PICU admission are shown for selected comorbidities compared with no comorbidity. Analyses within influenza admissions 2019/20 are not included due to low numbers.

**Figure S2** Odds ratios and percentage point difference in predicted probability with 95% confidence intervals for admission to PICU by body system comorbidities within each cohort, adjusted for sex, IMD category, ethnicity in 11-17 year olds only

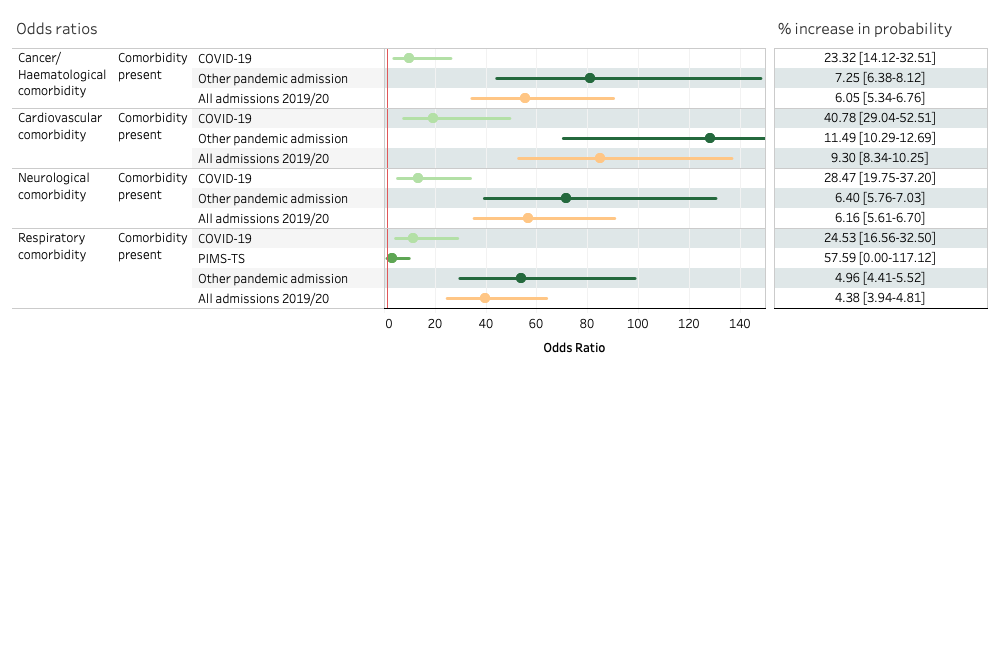

Notes: Results compare odds of PICU admission in selected comorbidity compared with no comorbidity. Upper limits for confidence intervals have been truncated for selected comorbidities for clarity. The red line indicates odds ratio of 1. Percentage point difference in predicted probability of PICU admission are shown for selected comorbidities compared with no comorbidity. Analyses within influenza admissions 2019/20 are not included due to low numbers.

**Figure S3** Odds ratios and predicted probability with 95% confidence intervals for admission to PICU by comorbidity combinations within each cohort, adjusted for sex, IMD category, ethnicity in 11-17 year olds only

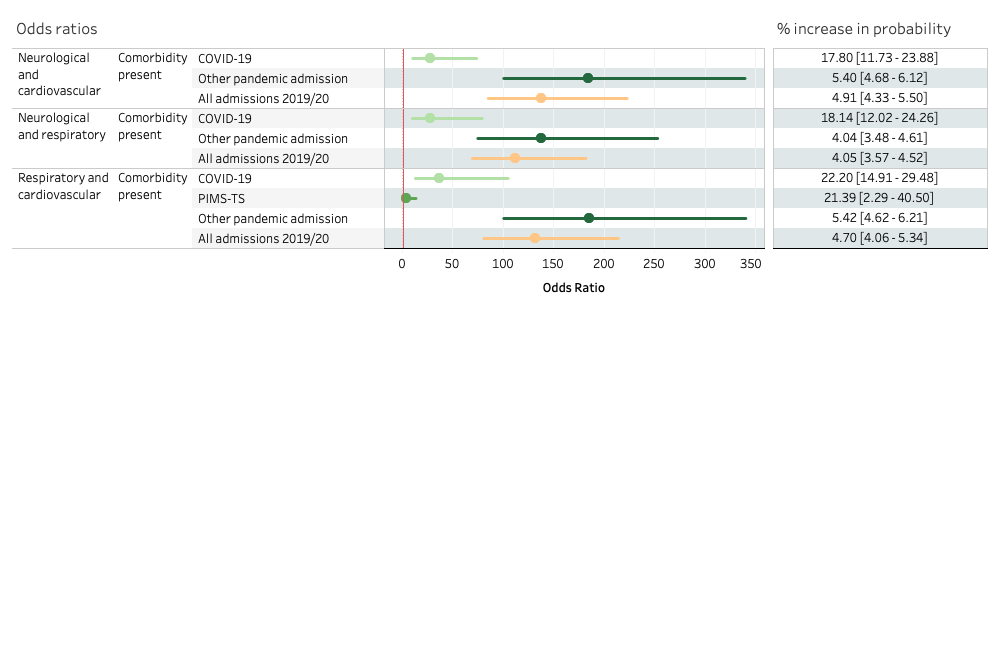

Notes: Results compare odds of PICU admission in selected comorbidity compared with no comorbidity. The red line indicates odds ratio of 1. Percentage point difference in predicted probability of PICU admission are shown for selected comorbidities compared with no comorbidity. Analyses within influenza admissions 2019/20 are not included due to low numbers.

**Figure S4** Odds ratios and predicted probability with 95% confidence intervals for admission to PICU by selected diagnoses within each cohort, adjusted for sex, IMD category, ethnicity in 11-17 year olds only

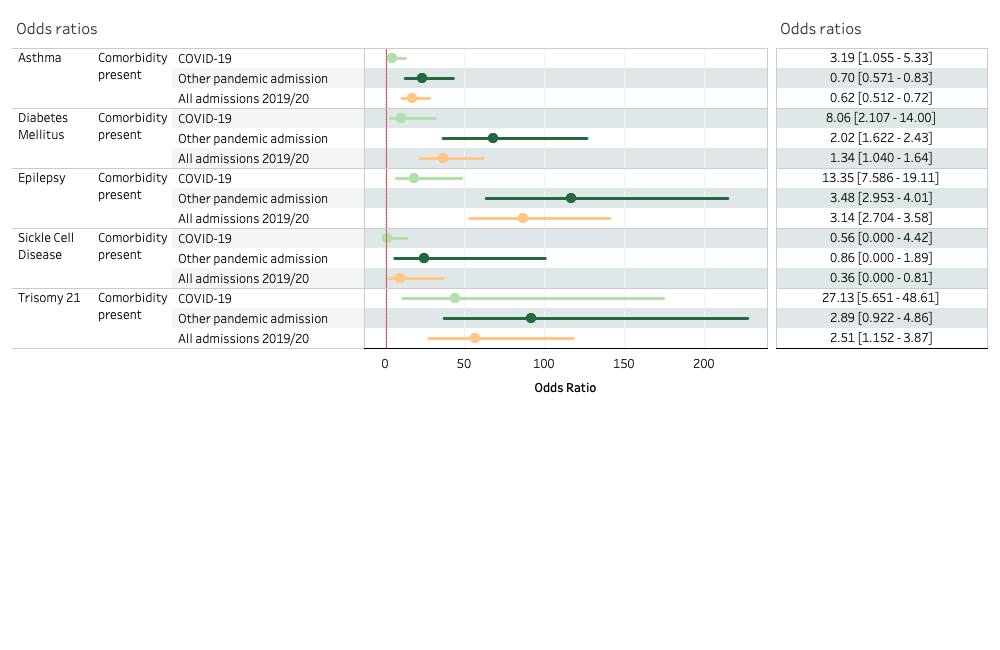

Notes: Results compare odds of PICU admission in selected comorbidity compared with no comorbidity. Upper limits for confidence intervals have been truncated for selected comorbidities for clarity. The red line indicates odds ratio of 1. Percentage point difference in predicted probability of PICU admission are shown for selected comorbidities compared with no comorbidity.

| **Table S16** Number of individual children and young people within each cohort by comorbidity group and paediatric intensive care admission | | | |
| --- | --- | --- | --- |
|  | All | | |
|  | Total | PICU | % PICU |
| COVID-19 | 5830 | 251 | 4.31 |
| PIMS-TS | 690 | 309 | 44.78 |
| Other pandemic year admission | 367637 | 4655 | 1.27 |
| All admissions 2019/20 | 587115 | 6621 | 1.13 |
| Influenza admission 2019/20 | 6780 | 160 | 2.36 |
|  | Any comorbidity | | |
|  | Total | PICU | % PICU |
| COVID-19 | 3020 | 229 | 7.58 |
| PIMS-TS | 435 | 259 | 59.54 |
| Other pandemic year admission | 153315 | 4301 | 2.81 |
| All admissions 2019/20 | 229546 | 5827 | 2.54 |
| Influenza admission 2019/20 | 2992 | 130 | 4.34 |
|  | Neurological and Respiratory | | |
|  | Total | PICU | % PICU |
| COVID-19 | 414 | 80 | 19.32 |
| PIMS-TS | 31 | 24 | 77.42 |
| Other pandemic year admission | 12575 | 1185 | 9.42 |
| All admissions 2019/20 | 19153 | 1843 | 9.62 |
| Influenza admission 2019/20 | 484 | 59 | 12.19 |
|  | Neurological and cardiovascular | | |
|  | Total | PICU | % PICU |
| COVID-19 | 441 | 93 | 21.09 |
| PIMS-TS | 66 | 49 | 74.24 |
| Other pandemic year admission | 11127 | 1463 | 13.15 |
| All admissions 2019/20 | 15276 | 1923 | 12.59 |
| Influenza admission 2019/20 | 399 | 37 | 9.27 |
|  | Respiratory and cardiovascular | | |
|  | Total | PICU | % PICU |
| COVID-19 | 354 | 80 | 22.60 |
| PIMS-TS | 66 | 49 | 74.24 |
| Other pandemic year admission | 9152 | 1254 | 13.70 |
| All admissions 2019/20 | 13311 | 1809 | 13.59 |
| Influenza admission 2019/20 | 406 | 44 | 10.84 |
|  | Cancer / Haematological | | |
|  | Total | PICU | % PICU |
| COVID-19 | 827 | 91 | 11.00 |
| PIMS-TS | 178 | 131 | 73.60 |
| Other pandemic year admission | 23192 | 1366 | 5.89 |
| All admissions 2019/20 | 32704 | 1809 | 5.53 |
| Influenza admission 2019/20 | 747 | 39 | 5.22 |
|  | Cardiovascular | | |
|  | Total | PICU | % PICU |
| COVID-19 | 863 | 150 | 17.38 |
| PIMS-TS | 275 | 177 | 64.36 |
| Other pandemic year admission | 24746 | 2611 | 10.55 |
| All admissions 2019/20 | 33863 | 3280 | 9.69 |
| Influenza admission 2019/20 | 781 | 55 | 7.04 |
|  | Neurological | | |
|  | Total | PICU | % PICU |
| COVID-19 | 1070 | 143 | 13.36 |
| PIMS-TS | 93 | 61 | 65.59 |
| Other pandemic year admission | 45273 | 2391 | 5.28 |
| All admissions 2019/20 | 65781 | 3250 | 4.94 |
| Influenza admission 2019/20 | 1047 | 84 | 8.02 |
|  | Respiratory | | |
|  | Total | PICU | % PICU |
| COVID-19 | 1020 | 114 | 11.18 |
| PIMS-TS | 100 | 65 | 65.00 |
| Other pandemic year admission | 46421 | 1971 | 4.25 |
| All admissions 2019/20 | 78321 | 3090 | 3.95 |
| Influenza admission 2019/20 | 1277 | 94 | 7.36 |
|  | Epilepsy | | |
|  | Total | PICU | % PICU |
| COVID-19 | 329 | 56 | 17.02 |
| PIMS-TS | 14 | 11 | 78.57 |
| Other pandemic year admission | 10193 | 781 | 7.66 |
| All admissions 2019/20 | 13977 | 1093 | 7.82 |
| Influenza admission 2019/20 | 302 | 44 | 14.57 |
|  | Asthma | | |
|  | Total | PICU | % PICU |
| COVID-19 | 545 | 30 | 5.50 |
| PIMS-TS | 47 | 26 | 55.32 |
| Other pandemic year admission | 29837 | 329 | 1.10 |
| All admissions 2019/20 | 51641 | 542 | 1.05 |
| Influenza admission 2019/20 | 664 | 15 | 2.26 |
|  | Trisomy 21 | | |
|  | Total | PICU | % PICU |
| COVID-19 | 60 | 16 | 26.67 |
| PIMS-TS | 1 | 0 | 0.00 |
| Other pandemic year admission | 1451 | 104 | 7.17 |
| All admissions 2019/20 | 2372 | 171 | 7.21 |
| Influenza admission 2019/20 | 47 | 4 | 8.51 |
|  | Diabetes | | |
|  | Total | PICU | % PICU |
| COVID-19 | 142 | 16 | 11.27 |
| PIMS-TS | 4 | 3 | 75.00 |
| Other pandemic year admission | 6471 | 199 | 3.08 |
| All admissions 2019/20 | 7658 | 157 | 2.05 |
| Influenza admission 2019/20 | 49 | 1 | 2.04 |
|  | Sickle Cell Disease | | |
|  | Total | PICU | % PICU |
| COVID-19 | 88 | 3 | 3.41 |
| PIMS-TS | 8 | 7 | 87.50 |
| Other pandemic year admission | 1898 | 52 | 2.74 |
| All admissions 2019/20 | 2953 | 87 | 2.95 |
| Influenza admission 2019/20 | 68 | 1 | 1.47 |
