## Supplement 2 for "Risk factors for intensive care admission and death amongst children and young people admitted to hospital with COVID-19 and PIMS-TS in England during the first pandemic year"

| | exposurevalue | Cohort | observations | Odds Ratios | | | | Predicted probabilities | | | | |
|---|---|---|---|---|---|---|---|---|---|---|---|---|
| exposure | | | | value | lower | upper | p | | value | lower | upper | p |
| Any comorbidity | | COVID-19 | 6338 | 11.59 | 7.21 | 18.62 | 0.00 | | | | | |
| | | COVID-19 | | | | | | absolute value | 7.45 | 6.41 | 8.49 | 0.00 |
| | | COVID-19 | | | | | | absolute difference | 6.75 | 5.64 | 7.86 | 0.00 |
| | | PIMS-TS | 710 | 4.53 | 3.03 | 6.78 | 0.00 | | | | | |
| | | PIMS-TS | | | | | | absolute value | 53.86 | 49.50 | 58.22 | 0.00 |
| | | PIMS-TS | | | | | | absolute difference | 28.36 | 21.11 | 35.60 | 0.00 |
| | | Other pandemic year admissions | 463371 | 23.42 | 20.92 | 26.23 | 0.00 | | | | | |
| | | Other pandemic year admissions | | | | | | absolute value | 2.67 | 2.58 | 2.75 | 0.00 |
| | | Other pandemic year admissions | | | | | | absolute difference | 2.55 | 2.46 | 2.63 | 0.00 |
| | | All admissions 2019/20 | 771357 | 13.78 | 12.76 | 14.88 | 0.00 | | | | | |
| | | All admissions 2019/20 | | | | | | absolute value | 2.17 | 2.11 | 2.23 | 0.00 |
| | | All admissions 2019/20 | | | | | | absolute difference | 2.01 | 1.95 | 2.07 | 0.00 |
| | | Influenza admissions 2019/20 | 6968 | 6.03 | 4.04 | 9.01 | 0.00 | | | | | |
| | | Influenza admissions 2019/20 | | | | | | absolute value | 4.38 | 3.63 | 5.13 | 0.00 |
| | | Influenza admissions 2019/20 | | | | | | absolute difference | 3.62 | 2.82 | 4.42 | 0.00 |
| Number of body systems | 1 body system | COVID-19 | 6338 | 4.14 | 2.34 | 7.34 | 0.00 | | | | | |
| | | COVID-19 | | | | | | absolute value | 2.70 | 1.79 | 3.61 | 0.00 |
| | | COVID-19 | | | | | | absolute difference | 2.03 | 1.06 | 3.00 | 0.00 |
| | More than 1 body system | COVID-19 | 6338 | 20.07 | 12.13 | 33.22 | 0.00 | | | | | |
| | | COVID-19 | | | | | | absolute value | 11.58 | 9.80 | 13.36 | 0.00 |
| | | COVID-19 | | | | | | absolute difference | 10.91 | 9.06 | 12.76 | 0.00 |
| | 1 body system | PIMS-TS | 710 | 2.59 | 1.61 | 4.15 | 0.00 | | | | | |
| | | PIMS-TS | | | | | | absolute value | 42.63 | 35.95 | 49.31 | 0.00 |
| | | PIMS-TS | | | | | | absolute difference | 17.39 | 8.72 | 26.06 | 0.00 |
| | More than 1 body system | PIMS-TS | 710 | 6.93 | 4.42 | 10.88 | 0.00 | | | | | |
| | | PIMS-TS | | | | | | absolute value | 62.14 | 56.41 | 67.87 | 0.00 |
| | | PIMS-TS | | | | | | absolute difference | 36.90 | 28.68 | 45.11 | 0.00 |
| | 1 body system | Other pandemic year admissions | 463371 | 9.66 | 8.52 | 10.96 | 0.00 | | | | | |
| | | Other pandemic year admissions | | | | | | absolute value | 1.10 | 1.04 | 1.17 | 0.00 |
| | | Other pandemic year admissions | | | | | | absolute difference | 0.99 | 0.92 | 1.06 | 0.00 |
| | More than 1 body system | Other pandemic year admissions | 463371 | 48.58 | 43.14 | 54.71 | 0.00 | | | | | |
| | | Other pandemic year admissions | | | | | | absolute value | 5.11 | 4.92 | 5.29 | 0.00 |
| | | Other pandemic year admissions | | | | | | absolute difference | 4.99 | 4.80 | 5.18 | 0.00 |
| | 1 body system | All admissions 2019/20 | 771357 | 5.73 | 5.24 | 6.27 | 0.00 | | | | | |
| | | All admissions 2019/20 | | | | | | absolute value | 0.91 | 0.86 | 0.96 | 0.00 |
| | | All admissions 2019/20 | | | | | | absolute difference | 0.75 | 0.70 | 0.80 | 0.00 |
| | More than 1 body system | All admissions 2019/20 | 771357 | 27.58 | 25.45 | 29.89 | 0.00 | | | | | |
| | | All admissions 2019/20 | | | | | | absolute value | 4.13 | 4.00 | 4.26 | 0.00 |
| | | All admissions 2019/20 | | | | | | absolute difference | 3.97 | 3.84 | 4.09 | 0.00 |
| | 1 body system | Influenza admissions 2019/20 | 6968 | 2.82 | 1.68 | 4.72 | 0.00 | | | | | |
| | | Influenza admissions 2019/20 | | | | | | absolute value | 2.09 | 1.34 | 2.84 | 0.00 |
| | | Influenza admissions 2019/20 | | | | | | absolute difference | 1.33 | 0.54 | 2.13 | 0.00 |
| | More than 1 body system | Influenza admissions 2019/20 | 6968 | 9.18 | 6.05 | 13.92 | 0.00 | | | | | |
| | | Influenza admissions 2019/20 | | | | | | absolute value | 6.45 | 5.21 | 7.70 | 0.00 |
| | | Influenza admissions 2019/20 | | | | | | absolute difference | 5.70 | 4.42 | 6.98 | 0.00 |
| niting or non life limiting como | Non life limiting comorbidity | COVID-19 | 6338 | 5.53 | 3.29 | 9.31 | 0.00 | | | | | |
| | | COVID-19 | | | | | | absolute value | 3.72 | 2.85 | 4.59 | 0.00 |
| | | COVID-19 | | | | | | absolute difference | 3.02 | 2.07 | 3.97 | 0.00 |
| | Life limiting comorbidity | COVID-19 | 6338 | 26.08 | 15.88 | 42.84 | 0.00 | | | | | |
| | | COVID-19 | | | | | | absolute value | 15.06 | 12.66 | 17.45 | 0.00 |
| | | COVID-19 | | | | | | absolute difference | 14.36 | 11.92 | 16.80 | 0.00 |
| | Non life limiting comorbidity | PIMS-TS | 710 | 3.69 | 2.41 | 5.64 | 0.00 | | | | | |
| | | PIMS-TS | | | | | | absolute value | 49.92 | 44.67 | 55.18 | 0.00 |
| | | PIMS-TS | | | | | | absolute difference | 24.49 | 16.69 | 32.28 | 0.00 |
| | Life limiting comorbidity | PIMS-TS | 710 | 7.18 | 4.20 | 12.27 | 0.00 | | | | | |
| | | PIMS-TS | | | | | | absolute value | 62.97 | 55.05 | 70.89 | 0.00 |
| | | PIMS-TS | | | | | | absolute difference | 37.54 | 27.70 | 47.38 | 0.00 |
| | Non life limiting comorbidity | Other pandemic year admissions | 463371 | 11.35 | 10.07 | 12.80 | 0.00 | | | | | |
| | | Other pandemic year admissions | | | | | | absolute value | 1.32 | 1.26 | 1.39 | 0.00 |
| | | Other pandemic year admissions | | | | | | absolute difference | 1.20 | 1.13 | 1.27 | 0.00 |
| | Life limiting comorbidity | Other pandemic year admissions | 463371 | 72.13 | 64.01 | 81.27 | 0.00 | | | | | |
| | | Other pandemic year admissions | | | | | | absolute value | 7.46 | 7.17 | 7.75 | 0.00 |
| | | Other pandemic year admissions | | | | | | absolute difference | 7.34 | 7.05 | 7.63 | 0.00 |
| | Non life limiting comorbidity | All admissions 2019/20 | 771357 | 6.47 | 5.95 | 7.04 | 0.00 | | | | | |
| | | All admissions 2019/20 | | | | | | absolute value | 1.04 | 1.00 | 1.09 | 0.00 |
| | | All admissions 2019/20 | | | | | | absolute difference | 0.88 | 0.83 | 0.93 | 0.00 |
| | Life limiting comorbidity | All admissions 2019/20 | 771357 | 43.96 | 40.53 | 47.67 | 0.00 | | | | | |
| | | All admissions 2019/20 | | | | | | absolute value | 6.45 | 6.24 | 6.65 | 0.00 |
| | | All admissions 2019/20 | | | | | | absolute difference | 6.28 | 6.07 | 6.49 | 0.00 |
| | Non life limiting comorbidity | Influenza admissions 2019/20 | 6968 | 2.92 | 1.82 | 4.68 | 0.00 | | | | | |
| | | Influenza admissions 2019/20 | | | | | | absolute value | 2.16 | 1.52 | 2.81 | 0.00 |
| | | Influenza admissions 2019/20 | | | | | | absolute difference | 1.41 | 0.71 | 2.11 | 0.00 |
| | Life limiting comorbidity | Influenza admissions 2019/20 | 6968 | 13.04 | 8.52 | 19.95 | 0.00 | | | | | |
| | | Influenza admissions 2019/20 | | | | | | absolute value | 8.89 | 7.07 | 10.71 | 0.00 |
| | | Influenza admissions 2019/20 | | | | | | absolute difference | 8.14 | 6.29 | 9.98 | 0.00 |
| Respiratory comorbidity | | COVID-19 | 4101 | 19.03 | 10.98 | 32.98 | 0.00 | | | | | |
| | | COVID-19 | | | | | | absolute value | 11.88 | 9.13 | 14.63 | 0.00 |
| | | COVID-19 | | | | | | absolute difference | 11.16 | 8.34 | 13.98 | 0.00 |
| | | PIMS-TS | 361 | 4.59 | 2.43 | 8.69 | 0.00 | | | | | |
| | | PIMS-TS | | | | | | absolute value | 50.25 | 40.12 | 60.38 | 0.00 |
| | | PIMS-TS | | | | | | absolute difference | 26.39 | 14.48 | 38.29 | 0.00 |
| | | Other pandemic year admissions | 312844 | 42.44 | 37.26 | 48.35 | 0.00 | | | | | |
| | | Other pandemic year admissions | | | | | | absolute value | 5.01 | 4.74 | 5.29 | 0.00 |
| | | Other pandemic year admissions | | | | | | absolute difference | 4.88 | 4.61 | 5.16 | 0.00 |
| | | All admissions 2019/20 | 548982 | 26.01 | 23.82 | 28.41 | 0.00 | | | | | |
| | | All admissions 2019/20 | | | | | | absolute value | 4.10 | 3.93 | 4.27 | 0.00 |
| | | All admissions 2019/20 | | | | | | absolute difference | 3.93 | 3.76 | 4.11 | 0.00 |
| | | Influenza admissions 2019/20 | 5187 | 12.65 | 8.06 | 19.83 | 0.00 | | | | | |
| | | Influenza admissions 2019/20 | | | | | | absolute value | 8.34 | 6.59 | 10.10 | 0.00 |
| | | Influenza admissions 2019/20 | | | | | | absolute difference | 7.61 | 5.82 | 9.40 | 0.00 |
| Cardiovascular comorbidity | | COVID-19 | 3961 | 27.10 | 16.48 | 44.59 | 0.00 | | | | | |
| | | COVID-19 | | | | | | absolute value | 16.17 | 13.42 | 18.91 | 0.00 |
| | | COVID-19 | | | | | | absolute difference | 15.43 | 12.64 | 18.22 | 0.00 |
| | | PIMS-TS | 542 | 6.36 | 4.02 | 10.06 | 0.00 | | | | | |
| | | PIMS-TS | | | | | | absolute value | 57.76 | 52.26 | 63.26 | 0.00 |
| | | PIMS-TS | | | | | | absolute difference | 33.42 | 25.50 | 41.34 | 0.00 |
| | | Other pandemic year admissions | 289180 | 65.72 | 58.35 | 74.03 | 0.00 | | | | | |
| | | Other pandemic year admissions | | | | | | absolute value | 7.91 | 7.60 | 8.22 | 0.00 |
| | | Other pandemic year admissions | | | | | | absolute difference | 7.77 | 7.46 | 8.08 | 0.00 |
| | | All admissions 2019/20 | 489665 | 39.48 | 36.37 | 42.85 | 0.00 | | | | | |
| | | All admissions 2019/20 | | | | | | absolute value | 6.50 | 6.27 | 6.72 | 0.00 |
| | | All admissions 2019/20 | | | | | | absolute difference | 6.31 | 6.09 | 6.54 | 0.00 |
| | | Influenza admissions 2019/20 | 4690 | 10.08 | 6.34 | 16.01 | 0.00 | | | | | |
| | | Influenza admissions 2019/20 | | | | | | absolute value | 7.06 | 5.23 | 8.89 | 0.00 |
| | | Influenza admissions 2019/20 | | | | | | absolute difference | 6.29 | 4.43 | 8.14 | 0.00 |
| Neurological comorbidity | | COVID-19 | 4171 | 20.97 | 12.52 | 35.11 | 0.00 | | | | | |
| | | COVID-19 | | | | | | absolute value | 13.04 | 10.58 | 15.51 | 0.00 |
| | | COVID-19 | | | | | | absolute difference | 12.31 | 9.78 | 14.84 | 0.00 |
| | | PIMS-TS | 351 | 4.93 | 2.58 | 9.41 | 0.00 | | | | | |
| | | PIMS-TS | | | | | | absolute value | 51.17 | 41.05 | 61.30 | 0.00 |
| | | PIMS-TS | | | | | | absolute difference | 27.47 | 15.74 | 39.21 | 0.00 |
| | | Other pandemic year admissions | 315855 | 39.70 | 35.16 | 44.82 | 0.00 | | | | | |
| | | Other pandemic year admissions | | | | | | absolute value | 4.90 | 4.67 | 5.12 | 0.00 |
| | | Other pandemic year admissions | | | | | | absolute difference | 4.76 | 4.54 | 4.99 | 0.00 |
| | | All admissions 2019/20 | 533799 | 23.13 | 21.34 | 25.08 | 0.00 | | | | | |
| | | All admissions 2019/20 | | | | | | absolute value | 3.93 | 3.79 | 4.08 | 0.00 |
| | | All admissions 2019/20 | | | | | | absolute difference | 3.75 | 3.61 | 3.90 | 0.00 |

| | | | | | | | | | | | |
|---|---|---|---|---|---|---|---|---|---|---|---|
| | Influenza admissions 2019/20 | 4955 | 11.94 | 7.76 | 18.39 | 0.00 | | | | | |
| | Influenza admissions 2019/20 | | | | | | absolute value | 8.24 | 6.49 | 9.99 | 0.00 |
| | Influenza admissions 2019/20 | | | | | | absolute difference | 7.48 | 5.70 | 9.25 | 0.00 |
| Cancer/Haematological comorbidity | COVID-19 | 3930 | 16.03 | 9.06 | 28.35 | 0.00 | | | | | |
| | COVID-19 | | | | | | absolute value | 10.43 | 7.70 | 13.16 | 0.00 |
| | COVID-19 | | | | | | absolute difference | 9.70 | 6.90 | 12.50 | 0.00 |
| | PIMS-TS | 441 | 7.64 | 4.55 | 12.82 | 0.00 | | | | | |
| | PIMS-TS | | | | | | absolute value | 63.25 | 55.99 | 70.50 | 0.00 |
| | PIMS-TS | | | | | | absolute difference | 37.84 | 28.19 | 47.49 | 0.00 |
| | Other pandemic year admissions | 286006 | 43.31 | 37.56 | 49.95 | 0.00 | | | | | |
| | Other pandemic year admissions | | | | | | absolute value | 5.43 | 5.04 | 5.83 | 0.00 |
| | Other pandemic year admissions | | | | | | absolute difference | 5.30 | 4.90 | 5.69 | 0.00 |
| | All admissions 2019/20 | 484299 | 27.85 | 25.31 | 30.63 | 0.00 | | | | | |
| | All admissions 2019/20 | | | | | | absolute value | 4.67 | 4.42 | 4.93 | 0.00 |
| | All admissions 2019/20 | | | | | | absolute difference | 4.49 | 4.24 | 4.75 | 0.00 |
| | Influenza admissions 2019/20 | 4654 | 8.13 | 4.82 | 13.71 | 0.00 | | | | | |
| | Influenza admissions 2019/20 | | | | | | absolute value | 5.72 | 3.82 | 7.61 | 0.00 |
| | Influenza admissions 2019/20 | | | | | | absolute difference | 4.96 | 3.04 | 6.88 | 0.00 |
| Neurological and cardiovascular | COVID-19 | 3456 | 35.87 | 20.57 | 62.58 | 0.00 | | | | | |
| | COVID-19 | | | | | | absolute value | 20.08 | 15.64 | 24.53 | 0.00 |
| | COVID-19 | | | | | | absolute difference | 19.34 | 14.84 | 23.83 | 0.00 |
| | PIMS-TS | 323 | 8.00 | 3.50 | 18.28 | 0.00 | | | | | |
| | PIMS-TS | | | | | | absolute value | 59.10 | 45.59 | 72.62 | 0.00 |
| | PIMS-TS | | | | | | absolute difference | 36.14 | 21.33 | 50.96 | 0.00 |
| | Other pandemic year admissions | 265479 | 80.64 | 70.57 | 92.15 | 0.00 | | | | | |
| | Other pandemic year admissions | | | | | | absolute value | 9.68 | 9.14 | 10.21 | 0.00 |
| | Other pandemic year admissions | | | | | | absolute difference | 9.54 | 9.00 | 10.07 | 0.00 |
| | All admissions 2019/20 | 454734 | 48.83 | 44.64 | 53.42 | 0.00 | | | | | |
| | All admissions 2019/20 | | | | | | absolute value | 8.03 | 7.66 | 8.40 | 0.00 |
| | All admissions 2019/20 | | | | | | absolute difference | 7.85 | 7.48 | 8.22 | 0.00 |
| | Influenza admissions 2019/20 | 4289 | 14.21 | 8.49 | 23.80 | 0.00 | | | | | |
| | Influenza admissions 2019/20 | | | | | | absolute value | 9.47 | 6.53 | 12.40 | 0.00 |
| | Influenza admissions 2019/20 | | | | | | absolute difference | 8.69 | 5.75 | 11.64 | 0.00 |
| Neurological and respiratory | COVID-19 | 3414 | 32.44 | 18.23 | 57.75 | 0.00 | | | | | |
| | COVID-19 | | | | | | absolute value | 19.00 | 14.11 | 23.89 | 0.00 |
| | COVID-19 | | | | | | absolute difference | 18.26 | 13.32 | 23.20 | 0.00 |
| | PIMS-TS | 288 | 10.51 | 3.24 | 34.10 | 0.00 | | | | | |
| | PIMS-TS | | | | | | absolute value | 60.57 | 40.42 | 80.72 | 0.00 |
| | PIMS-TS | | | | | | absolute difference | 39.12 | 18.38 | 59.87 | 0.00 |
| | Other pandemic year admissions | 266108 | 68.90 | 59.63 | 79.61 | 0.00 | | | | | |
| | Other pandemic year admissions | | | | | | absolute value | 8.44 | 7.85 | 9.03 | 0.00 |
| | Other pandemic year admissions | | | | | | absolute difference | 8.30 | 7.70 | 8.89 | 0.00 |
| | All admissions 2019/20 | 459563 | 45.44 | 41.39 | 49.89 | 0.00 | | | | | |
| | All admissions 2019/20 | | | | | | absolute value | 7.47 | 7.09 | 7.85 | 0.00 |
| | All admissions 2019/20 | | | | | | absolute difference | 7.28 | 6.90 | 7.66 | 0.00 |
| | Influenza admissions 2019/20 | 4368 | 21.15 | 12.84 | 34.85 | 0.00 | | | | | |
| | Influenza admissions 2019/20 | | | | | | absolute value | 13.12 | 9.79 | 16.46 | 0.00 |
| | Influenza admissions 2019/20 | | | | | | absolute difference | 12.36 | 9.01 | 15.71 | 0.00 |
| Respiratory and cardiovascular | COVID-19 | 3357 | 37.74 | 21.65 | 65.80 | 0.00 | | | | | |
| | COVID-19 | | | | | | absolute value | 21.40 | 16.25 | 26.55 | 0.00 |
| | COVID-19 | | | | | | absolute difference | 20.65 | 15.47 | 25.84 | 0.00 |
| | PIMS-TS | 325 | 6.63 | 3.15 | 13.96 | 0.00 | | | | | |
| | PIMS-TS | | | | | | absolute value | 52.60 | 40.04 | 65.16 | 0.00 |
| | PIMS-TS | | | | | | absolute difference | 30.98 | 17.37 | 44.59 | 0.00 |
| | Other pandemic year admissions | 261424 | 85.24 | 74.19 | 97.95 | 0.00 | | | | | |
| | Other pandemic year admissions | | | | | | absolute value | 10.20 | 9.58 | 10.82 | 0.00 |
| | Other pandemic year admissions | | | | | | absolute difference | 10.06 | 9.44 | 10.68 | 0.00 |
| | All admissions 2019/20 | 450099 | 54.69 | 49.85 | 60.01 | 0.00 | | | | | |
| | All admissions 2019/20 | | | | | | absolute value | 8.83 | 8.41 | 9.25 | 0.00 |
| | All admissions 2019/20 | | | | | | absolute difference | 8.64 | 8.23 | 9.06 | 0.00 |
| | Influenza admissions 2019/20 | 4293 | 17.06 | 10.22 | 28.50 | 0.00 | | | | | |
| | Influenza admissions 2019/20 | | | | | | absolute value | 11.06 | 7.84 | 14.29 | 0.00 |
| | Influenza admissions 2019/20 | | | | | | absolute difference | 10.29 | 7.05 | 13.53 | 0.00 |
| Epilepsy | COVID-19 | 3316 | 22.99 | 12.47 | 42.39 | 0.00 | | | | | |
| | COVID-19 | | | | | | absolute value | 14.43 | 9.58 | 19.29 | 0.00 |
| | COVID-19 | | | | | | absolute difference | 13.67 | 8.77 | 18.58 | 0.00 |
| | PIMS-TS | 270 | 21.17 | 2.98 | 150.40 | 0.00 | | | | | |
| | PIMS-TS | | | | | | absolute value | 70.17 | 40.48 | 99.86 | 0.00 |
| | PIMS-TS | | | | | | absolute difference | 50.43 | 20.37 | 80.50 | 0.00 |
| | Other pandemic year admissions | 262103 | 50.88 | 43.85 | 59.04 | 0.00 | | | | | |
| | Other pandemic year admissions | | | | | | absolute value | 6.62 | 6.00 | 7.24 | 0.00 |
| | Other pandemic year admissions | | | | | | absolute difference | 6.48 | 5.86 | 7.10 | 0.00 |
| | All admissions 2019/20 | 448138 | 38.26 | 34.55 | 42.37 | 0.00 | | | | | |
| | All admissions 2019/20 | | | | | | absolute value | 6.50 | 6.05 | 6.94 | 0.00 |
| | All admissions 2019/20 | | | | | | absolute difference | 6.31 | 5.86 | 6.76 | 0.00 |
| | Influenza admissions 2019/20 | 4177 | 25.55 | 14.91 | 43.79 | 0.00 | | | | | |
| | Influenza admissions 2019/20 | | | | | | absolute value | 15.62 | 10.93 | 20.30 | 0.00 |
| | Influenza admissions 2019/20 | | | | | | absolute difference | 14.85 | 10.15 | 19.55 | 0.00 |
| Diabetes Mellitus | COVID-19 | 2951 | 17.97 | 6.91 | 46.73 | 0.00 | | | | | |
| | COVID-19 | | | | | | absolute value | 11.74 | 3.51 | 19.97 | 0.01 |
| | COVID-19 | | | | | | absolute difference | 10.96 | 2.69 | 19.22 | 0.01 |
| | PIMS-TS | 260 | 14.55 | 0.69 | 306.30 | 0.09 | | | | | |
| | PIMS-TS | | | | | | absolute value | 62.21 | 10.55 | 100* | 0.02 |
| | PIMS-TS | | | | | | absolute difference | 42.56 | -9.24 | 94.35 | 0.11 |
| | Other pandemic year admissions | 250458 | 45.32 | 35.79 | 57.40 | 0.00 | | | | | |
| | Other pandemic year admissions | | | | | | absolute value | 5.98 | 4.82 | 7.14 | 0.00 |
| | Other pandemic year admissions | | | | | | absolute difference | 5.84 | 4.68 | 7.00 | 0.00 |
| | All admissions 2019/20 | 428571 | 23.43 | 18.94 | 28.99 | 0.00 | | | | | |
| | All admissions 2019/20 | | | | | | absolute value | 4.13 | 3.35 | 4.91 | 0.00 |
| | All admissions 2019/20 | | | | | | absolute difference | 3.94 | 3.16 | 4.72 | 0.00 |
| | Influenza admissions 2019/20 | 3718 | 4.18 | 0.57 | 30.58 | 0.16 | | | | | |
| | Influenza admissions 2019/20 | | | | | | absolute value | 3.26 | 0.0* | 9.31 | 0.29 |
| | Influenza admissions 2019/20 | | | | | | absolute difference | 2.44 | -3.61 | 8.50 | 0.43 |
| Asthma | COVID-19 | 3541 | 7.48 | 3.49 | 16.01 | 0.00 | | | | | |
| | COVID-19 | | | | | | absolute value | 5.25 | 2.50 | 8.00 | 0.00 |
| | COVID-19 | | | | | | absolute difference | 4.50 | 1.67 | 7.33 | 0.00 |
| | PIMS-TS | 305 | 1.92 | 0.88 | 4.17 | 0.10 | | | | | |
| | PIMS-TS | | | | | | absolute value | 32.14 | 21.61 | 42.68 | 0.00 |
| | PIMS-TS | | | | | | absolute difference | 9.32 | -2.54 | 21.18 | 0.12 |
| | Other pandemic year admissions | 282815 | 11.39 | 9.24 | 14.05 | 0.00 | | | | | |
| | Other pandemic year admissions | | | | | | absolute value | 1.51 | 1.25 | 1.77 | 0.00 |
| | Other pandemic year admissions | | | | | | absolute difference | 1.37 | 1.12 | 1.63 | 0.00 |
| | All admissions 2019/20 | 494995 | 7.97 | 6.91 | 9.20 | 0.00 | | | | | |
| | All admissions 2019/20 | | | | | | absolute value | 1.37 | 1.21 | 1.53 | 0.00 |
| | All admissions 2019/20 | | | | | | absolute difference | 1.19 | 1.03 | 1.35 | 0.00 |
| | Influenza admissions 2019/20 | 4540 | 4.18 | 1.95 | 8.94 | 0.00 | | | | | |
| | Influenza admissions 2019/20 | | | | | | absolute value | 3.00 | 1.18 | 4.82 | 0.00 |
| | Influenza admissions 2019/20 | | | | | | absolute difference | 2.26 | 0.40 | 4.12 | 0.02 |
| Sickle Cell Disease | COVID-19 | 2891 | 1.07 | 0.25 | 4.48 | 0.93 | | | | | |
| | COVID-19 | | | | | | absolute value | 0.88 | 0.0* | 2.01 | 0.13 |
| | COVID-19 | | | | | | absolute difference | 0.05 | -1.16 | 1.27 | 0.93 |
| | PIMS-TS | 262 | 9.45 | 0.44 | 202.10 | 0.15 | | | | | |
| | PIMS-TS | | | | | | absolute value | 56.02 | 0.98 | 100* | 0.05 |
| | PIMS-TS | | | | | | absolute difference | 36.15 | -19.02 | 91.32 | 0.20 |
| | Other pandemic year admissions | 244932 | 11.84 | 7.89 | 17.77 | 0.00 | | | | | |
| | Other pandemic year admissions | | | | | | absolute value | 1.70 | 1.05 | 2.35 | 0.00 |
| | Other pandemic year admissions | | | | | | absolute difference | 1.55 | 0.90 | 2.20 | 0.00 |
| | All admissions 2019/20 | 422527 | 15.36 | 11.02 | 21.40 | 0.00 | | | | | |
| | All admissions 2019/20 | | | | | | absolute value | 2.80 | 1.93 | 3.67 | 0.00 |

| | | | | | | | | | | | |
|---|---|---|---|---|---|---|---|---|---|---|---|
| | All admissions 2019/20 | | | | | | absolute difference | 2.61 | 1.74 | 3.48 | 0.00 |
| | Influenza admissions 2019/20 | 3733 | 1.41 | 0.17 | 11.68 | 0.75 | | | | | |
| | Influenza admissions 2019/20 | | | | | | absolute value | 1.16 | 0.0* | 3.51 | 0.34 |
| | Influenza admissions 2019/20 | | | | | | absolute difference | 0.33 | -2.04 | 2.71 | 0.78 |
| Trisomy 21 | COVID-19 | 2990 | 34.80 | 15.75 | 76.89 | 0.00 | | | | | |
| | COVID-19 | | | | | | absolute value | 19.37 | 9.03 | 29.72 | 0.00 |
| | COVID-19 | | | | | | absolute difference | 18.61 | 8.26 | 28.96 | 0.00 |
| | Other pandemic year admissions | 244428 | 35.63 | 28.18 | 45.05 | 0.00 | | | | | |
| | Other pandemic year admissions | | | | | | absolute value | 4.87 | 3.92 | 5.81 | 0.00 |
| | Other pandemic year admissions | | | | | | absolute difference | 4.72 | 3.78 | 5.67 | 0.00 |
| | All admissions 2019/20 | 422074 | 29.92 | 25.02 | 35.77 | 0.00 | | | | | |
| | All admissions 2019/20 | | | | | | absolute value | 5.26 | 4.46 | 6.06 | 0.00 |
| | All admissions 2019/20 | | | | | | absolute difference | 5.07 | 4.27 | 5.87 | 0.00 |
| | Influenza admissions 2019/20 | 3723 | 10.83 | 3.60 | 32.58 | 0.00 | | | | | |
| | Influenza admissions 2019/20 | | | | | | absolute value | 7.83 | 0.71 | 14.96 | 0.03 |
| | Influenza admissions 2019/20 | | | | | | absolute difference | 7.02 | -0.12 | 14.15 | 0.05 |

*Confidence intervals for abosolute predicted probability of PICU admission which exceed 100 or are lower than 0 are rounded to 100 and 0 respectively

Data are odds ratios with confidence intervals multivariable GEE models adjusted
Predicted probabilities show absolute and relative differnence in predicted probal

l for age, sex, ethnicity and IMD
bility of PICU admission by comorbidity group compared with no comorbidity
