## Supplement 3 for "Risk factors for intensive care admission and death amongst children and young people admitted to hospital with COVID-19 and PIMS-TS in England during the first pandemic year"

| exposure | | | Odds ratios from GEE models | | | | percentage change in relative predicted probability (%) | | | | abslute predicted probability (%) | | | | |
|---|---|---|---|---|---|---|---|---|---|---|---|---|---|---|---|
| | | cohort | OR | lower | upper | p | relative_change | lower | upper | p | absolute_probability | lower | upper | p | Age group |
| Any comorbidity | | COVID-19 | 6.30 | 2.54 | 15.64 | 0.00 | 4.61 | 3.20 | 6.03 | 0.00 | 5.56 | 4.41 | 6.70 | 0.00 | 11-17 |
| | | PIMS-TS | 4.98 | 2.04 | 12.18 | 0.00 | 31.14 | 13.55 | 48.74 | 0.00 | 78.51 | 72.04 | 84.98 | 0.00 | 11-17 |
| | | Other pandemic admission | 33.24 | 18.31 | 60.35 | 0.00 | 0.97 | 0.90 | 1.05 | 0.00 | 1.01 | 0.93 | 1.08 | 0.00 | 11-17 |
| | | All admissions 2019/20 | 23.16 | 14.50 | 37.02 | 0.00 | 0.82 | 0.76 | 0.89 | 0.00 | 0.86 | 0.80 | 0.92 | 0.00 | 11-17 |
| | | Influenza admissions 2019/20 | - | - | - | - | | | | | 3.83 | 2.36 | 5.30 | 0.00 | 11-17 |
| Asthma | | COVID-19 | 4.72 | 1.74 | 12.85 | 0.00 | 3.19 | 1.05 | 5.33 | 0.00 | 4.11 | 2.09 | 6.12 | 0.00 | 11-17 |
| | | Other pandemic admission | 23.20 | 12.56 | 42.84 | 0.00 | 0.70 | 0.57 | 0.83 | 0.00 | 0.73 | 0.60 | 0.86 | 0.00 | 11-17 |
| | | All admissions 2019/20 | 17.11 | 10.48 | 27.94 | 0.00 | 0.62 | 0.51 | 0.72 | 0.00 | 0.66 | 0.55 | 0.76 | 0.00 | 11-17 |
| | | Influenza admissions 2019/20 | - | - | - | - | | | | | 3.45 | 0.74 | 6.16 | 0.01 | 11-17 |
| Cancer/Haematological comorbidity | | COVID-19 | 9.82 | 3.69 | 26.16 | 0.00 | 7.77 | 4.71 | 10.84 | 0.00 | 8.75 | 5.85 | 11.64 | 0.00 | 11-17 |
| | | Other pandemic admission | 80.93 | 44.19 | 148.20 | 0.00 | 2.42 | 2.13 | 2.71 | 0.00 | 2.45 | 2.16 | 2.74 | 0.00 | 11-17 |
| | | All admissions 2019/20 | 55.70 | 34.40 | 90.18 | 0.00 | 2.02 | 1.78 | 2.25 | 0.00 | 2.05 | 1.82 | 2.29 | 0.00 | 11-17 |
| | | Influenza admissions 2019/20 | - | - | - | - | | | | | 5.17 | 1.71 | 8.63 | 0.00 | 11-17 |
| Cardiovascular comorbidity | | COVID-19 | 19.29 | 7.54 | 49.37 | 0.00 | 13.59 | 9.68 | 17.50 | 0.00 | 14.50 | 10.69 | 18.32 | 0.00 | 11-17 |
| | | Other pandemic admission | 128.40 | 70.41 | 234.30 | 0.00 | 3.83 | 3.43 | 4.23 | 0.00 | 3.86 | 3.46 | 4.26 | 0.00 | 11-17 |
| | | All admissions 2019/20 | 84.98 | 52.81 | 136.80 | 0.00 | 3.10 | 2.78 | 3.42 | 0.00 | 3.14 | 2.82 | 3.45 | 0.00 | 11-17 |
| | | Influenza admissions 2019/20 | - | - | - | - | | | | | 10.15 | 3.51 | 16.79 | 0.00 | 11-17 |
| Diabetes Mellitus | | COVID-19 | 10.38 | 3.42 | 31.58 | 0.00 | 8.06 | 2.11 | 14.00 | 0.01 | 9.05 | 3.13 | 14.97 | 0.00 | 11-17 |
| | | Other pandemic admission | 67.74 | 36.29 | 126.40 | 0.00 | 2.02 | 1.62 | 2.43 | 0.00 | 2.06 | 1.65 | 2.46 | 0.00 | 11-17 |
| | | All admissions 2019/20 | 36.72 | 21.95 | 61.43 | 0.00 | 1.34 | 1.04 | 1.64 | 0.00 | 1.38 | 1.08 | 1.68 | 0.00 | 11-17 |
| Epilepsy | | COVID-19 | 18.44 | 7.08 | 48.04 | 0.00 | 13.35 | 7.59 | 19.11 | 0.00 | 14.32 | 8.56 | 20.07 | 0.00 | 11-17 |
| | | Other pandemic admission | 116.50 | 63.29 | 214.50 | 0.00 | 3.48 | 2.95 | 4.01 | 0.00 | 3.51 | 2.98 | 4.04 | 0.00 | 11-17 |
| | | All admissions 2019/20 | 86.37 | 52.99 | 140.80 | 0.00 | 3.14 | 2.70 | 3.58 | 0.00 | 3.18 | 2.74 | 3.62 | 0.00 | 11-17 |
| | | Influenza admissions 2019/20 | - | - | - | - | | | | | 15.80 | 7.61 | 23.99 | 0.00 | 11-17 |
| Life limiting neurodisability | | COVID-19 | 26.63 | 7.93 | 89.48 | 0.00 | 17.00 | 4.79 | 29.21 | 0.01 | 17.88 | 5.66 | 30.10 | 0.00 | 11-17 |
| | | Other pandemic admission | 187.50 | 100.60 | 349.20 | 0.00 | 5.44 | 4.24 | 6.65 | 0.00 | 5.47 | 4.27 | 6.68 | 0.00 | 11-17 |
| | | All admissions 2019/20 | 137.90 | 82.65 | 230.00 | 0.00 | 4.91 | 3.91 | 5.91 | 0.00 | 4.94 | 3.94 | 5.94 | 0.00 | 11-17 |
| Life limiting or non life limiting comorbidity | Life limiting | COVID-19 | 16.55 | 6.54 | 41.92 | 0.00 | 12.19 | 8.75 | 15.62 | 0.00 | 13.13 | 9.79 | 16.46 | 0.00 | 11-17 |
| | | PIMS-TS | 4.17 | 1.48 | 11.75 | 0.01 | 28.25 | 8.83 | 47.68 | 0.00 | 75.51 | 64.03 | 86.99 | 0.00 | 11-17 |
| | | Other pandemic admission | 135.30 | 74.35 | 246.30 | 0.00 | 3.94 | 3.57 | 4.32 | 0.00 | 3.97 | 3.60 | 4.35 | 0.00 | 11-17 |
| | | All admissions 2019/20 | 96.65 | 60.30 | 154.90 | 0.00 | 3.47 | 3.16 | 3.78 | 0.00 | 3.50 | 3.20 | 3.81 | 0.00 | 11-17 |
| | | Influenza admissions 2019/20 | - | - | - | - | | | | | 9.80 | 5.61 | 13.99 | 0.00 | 11-17 |
| | Non life limiting | COVID-19 | 3.06 | 1.18 | 7.91 | 0.02 | 1.86 | 0.60 | 3.13 | 0.00 | 2.81 | 1.84 | 3.77 | 0.00 | 11-17 |
| | | PIMS-TS | 5.57 | 2.12 | 14.68 | 0.00 | 32.77 | 14.25 | 51.29 | 0.00 | 80.03 | 72.10 | 87.95 | 0.00 | 11-17 |
| | | Other pandemic admission | 14.43 | 7.89 | 26.41 | 0.00 | 0.41 | 0.35 | 0.47 | 0.00 | 0.44 | 0.39 | 0.49 | 0.00 | 11-17 |
| | | All admissions 2019/20 | 9.55 | 5.93 | 15.40 | 0.00 | 0.32 | 0.28 | 0.37 | 0.00 | 0.36 | 0.32 | 0.40 | 0.00 | 11-17 |
| | | Influenza admissions 2019/20 | 0.12 | 0.05 | 0.31 | 0.00 | | | | | 1.31 | 0.27 | 2.34 | 0.01 | 11-17 |
| Neurological and cardiovascular | | COVID-19 | 27.68 | 10.38 | 73.79 | 0.00 | 17.80 | 11.73 | 23.88 | 0.00 | 18.69 | 12.68 | 24.71 | 0.00 | 11-17 |
| | | Other pandemic admission | 184.80 | 100.60 | 339.50 | 0.00 | 5.40 | 4.68 | 6.12 | 0.00 | 5.43 | 4.71 | 6.15 | 0.00 | 11-17 |
| | | All admissions 2019/20 | 137.80 | 85.22 | 222.80 | 0.00 | 4.91 | 4.33 | 5.50 | 0.00 | 4.95 | 4.37 | 5.54 | 0.00 | 11-17 |
| | | Influenza admissions 2019/20 | - | - | - | - | | | | | 11.45 | 1.24 | 21.66 | 0.03 | 11-17 |
| Neurological and respiratory | | COVID-19 | 28.38 | 10.15 | 79.34 | 0.00 | 18.14 | 12.02 | 24.26 | 0.00 | 19.02 | 12.99 | 25.05 | 0.00 | 11-17 |
| | | Other pandemic admission | 137.70 | 74.91 | 253.00 | 0.00 | 4.04 | 3.48 | 4.61 | 0.00 | 4.07 | 3.51 | 4.64 | 0.00 | 11-17 |
| | | All admissions 2019/20 | 112.50 | 69.48 | 182.00 | 0.00 | 4.05 | 3.57 | 4.52 | 0.00 | 4.08 | 3.61 | 4.56 | 0.00 | 11-17 |
| | | Influenza admissions 2019/20 | - | - | - | - | | | | | 14.57 | 7.59 | 21.55 | 0.00 | 11-17 |
| Neurological comorbidity | | COVID-19 | 13.19 | 5.14 | 33.87 | 0.00 | 9.49 | 6.58 | 12.40 | 0.00 | 10.40 | 7.62 | 13.18 | 0.00 | 11-17 |
| | | Other pandemic admission | 71.65 | 39.33 | 130.50 | 0.00 | 2.13 | 1.92 | 2.34 | 0.00 | 2.16 | 1.95 | 2.37 | 0.00 | 11-17 |
| | | All admissions 2019/20 | 56.57 | 35.29 | 90.66 | 0.00 | 2.05 | 1.87 | 2.23 | 0.00 | 2.09 | 1.91 | 2.27 | 0.00 | 11-17 |
| | | Influenza admissions 2019/20 | - | - | - | - | | | | | 8.60 | 4.83 | 12.37 | 0.00 | 11-17 |
| Number of body systems | 1 body system | COVID-19 | 1.50 | 0.49 | 4.58 | 0.48 | 0.46 | 0.00 | 1.70 | 0.47 | 1.39 | 0.44 | 2.35 | 0.00 | 11-17 |
| | | PIMS-TS | 4.18 | 1.43 | 12.23 | 0.01 | 28.40 | 7.86 | 48.94 | 0.01 | 75.53 | 63.28 | 87.77 | 0.00 | 11-17 |
| | | Other pandemic admission | 11.23 | 6.05 | 20.84 | 0.00 | 0.31 | 0.25 | 0.37 | 0.00 | 0.34 | 0.28 | 0.40 | 0.00 | 11-17 |
| | | All admissions 2019/20 | 7.27 | 4.43 | 11.93 | 0.00 | 0.23 | 0.18 | 0.28 | 0.00 | 0.27 | 0.22 | 0.32 | 0.00 | 11-17 |
| | | Influenza admissions 2019/20 | 0.13 | 0.04 | 0.44 | 0.00 | | | | | 0.94 | 0.00 | 1.98 | 0.08 | 11-17 |
| | More than 1 body system | COVID-19 | 9.65 | 3.86 | 24.15 | 0.00 | 7.22 | 5.26 | 9.18 | 0.00 | 8.16 | 6.38 | 9.93 | 0.00 | 11-17 |
| | | PIMS-TS | 5.56 | 2.16 | 14.30 | 0.00 | 32.88 | 14.96 | 50.79 | 0.00 | 80.01 | 72.36 | 87.65 | 0.00 | 11-17 |
| | | Other pandemic admission | 57.26 | 31.50 | 104.10 | 0.00 | 1.67 | 1.53 | 1.81 | 0.00 | 1.70 | 1.56 | 1.84 | 0.00 | 11-17 |
| | | All admissions 2019/20 | 40.30 | 25.19 | 64.48 | 0.00 | 1.43 | 1.32 | 1.55 | 0.00 | 1.47 | 1.36 | 1.58 | 0.00 | 11-17 |
| | | Influenza admissions 2019/20 | | | | | | | | | 6.59 | 3.94 | 9.24 | 0.00 | 11-17 |
| Respiratory and cardiovascular | | COVID-19 | 37.07 | 13.09 | 105.00 | 0.00 | 22.20 | 14.91 | 29.48 | 0.00 | 23.07 | 15.86 | 30.27 | 0.00 | 11-17 |
| | | PIMS-TS | 4.06 | 1.20 | 13.70 | 0.02 | 21.39 | 2.29 | 40.50 | 0.03 | 75.19 | 63.80 | 86.59 | 0.00 | 11-17 |
| | | Other pandemic admission | 185.10 | 100.60 | 340.30 | 0.00 | 5.42 | 4.62 | 6.21 | 0.00 | 5.45 | 4.65 | 6.25 | 0.00 | 11-17 |
| | | All admissions 2019/20 | 131.50 | 80.78 | 214.10 | 0.00 | 4.70 | 4.06 | 5.34 | 0.00 | 4.74 | 4.10 | 5.38 | 0.00 | 11-17 |
| | | Influenza admissions 2019/20 | - | - | - | - | | | | | 22.39 | 6.47 | 38.32 | 0.01 | 11-17 |
| Respiratory comorbidity | | COVID-19 | 11.23 | 4.37 | 28.83 | 0.00 | 8.18 | 5.52 | 10.83 | 0.00 | 9.08 | 6.55 | 11.61 | 0.00 | 11-17 |
| | | PIMS-TS | 3.11 | 0.99 | 9.74 | 0.05 | 19.20 | 0.00 | 39.04 | 0.06 | 72.01 | 59.54 | 84.48 | 0.00 | 11-17 |
| | | Other pandemic admission | 54.11 | 29.68 | 98.66 | 0.00 | 1.65 | 1.47 | 1.84 | 0.00 | 1.69 | 1.50 | 1.87 | 0.00 | 11-17 |

| | | | | | | | | | | | | | | |
|---|---|---|---|---|---|---|---|---|---|---|---|---|---|---|
| | All admissions 2019/20 | 39.70 | 24.68 | 63.84 | 0.00 | 1.46 | 1.31 | 1.60 | 0.00 | 1.50 | 1.35 | 1.64 | 0.00 | 11-17 |
| | Influenza admissions 2019/20 | - | - | - | - | | | | | 6.53 | 3.77 | 9.30 | 0.00 | 11-17 |
| Sickle Cell Disease | COVID-19 | 1.44 | 0.15 | 13.71 | 0.75 | 0.56 | 0.00 | 4.42 | 0.77 | 1.93 | 0.00 | 5.29 | 0.26 | 11-17 |
| | Other pandemic admission | 24.65 | 6.06 | 100.30 | 0.00 | 0.86 | 0.00 | 1.89 | 0.10 | 0.90 | 0.00 | 1.92 | 0.09 | 11-17 |
| | All admissions 2019/20 | 9.44 | 2.46 | 36.18 | 0.00 | 0.36 | 0.00 | 0.81 | 0.11 | 0.41 | 0.00 | 0.85 | 0.07 | 11-17 |
| Trisomy 21 | COVID-19 | 43.83 | 11.02 | 174.40 | 0.00 | 27.13 | 5.65 | 48.61 | 0.01 | 28.10 | 6.64 | 49.57 | 0.01 | 11-17 |
| | Other pandemic admission | 91.61 | 36.97 | 227.00 | 0.00 | 2.89 | 0.92 | 4.86 | 0.00 | 2.92 | 0.95 | 4.89 | 0.00 | 11-17 |
| | All admissions 2019/20 | 56.84 | 27.40 | 117.90 | 0.00 | 2.51 | 1.15 | 3.87 | 0.00 | 2.56 | 1.20 | 3.92 | 0.00 | 11-17 |

*Confidence intervals for abosolute predicted probability of PICU admission which exceed 100 or are lower than 0 are rounded to 100 and 0 respectively
Where models did not run due to low numbers within specific cohorts these are excluded

Odds ratios with 95% confidence intervals for admission to PICU by number of comorbidities in 11-17 year olds only (full gee models)

| Cohort | exposure | exposurevalue | OR | lower | upper | p_value | Observations |
|---|---|---|---|---|---|---|---|
| COVID-19 | Sex | Female | 0.62 | 0.40 | 0.97 | 0.04 | 2046 |
| | IMD | 2nd most deprived quintil | 1.33 | 0.75 | 2.34 | 0.33 | |
| | IMD | 3rd most deprived quintil | 0.99 | 0.50 | 1.95 | 0.97 | |
| | IMD | 4th most deprived quintil | 0.69 | 0.30 | 1.60 | 0.39 | |
| | IMD | Least deprived quintile | 1.38 | 0.63 | 3.04 | 0.42 | |
| | Ethnicity | Mixed | 0.38 | 0.05 | 2.92 | 0.36 | |
| | Ethnicity | Asian | 1.36 | 0.74 | 2.51 | 0.32 | |
| | Ethnicity | Black | 3.17 | 1.69 | 5.94 | 0.00 | |
| | Ethnicity | Other | 0.75 | 0.22 | 2.59 | 0.65 | |
| | Ethnicity | Unknown | 1.26 | 0.54 | 2.90 | 0.60 | |
| | Number of body systems | Comorbidity present | 1.50 | 0.49 | 4.58 | 0.48 | |
| | Number of body systems | More than 1 body system | 9.65 | 3.86 | 24.15 | 0.00 | |
| PIMS-TS | Sex | Female | 1.36 | 0.63 | 2.97 | 0.43 | 173 |
| | IMD | 2nd most deprived quintil | 1.28 | 0.49 | 3.35 | 0.61 | |
| | IMD | 3rd most deprived quintil | 7.32 | 1.67 | 32.17 | 0.01 | |
| | IMD | 4th most deprived quintil | 6.03 | 1.35 | 26.97 | 0.02 | |
| | IMD | Least deprived quintile | 2.14 | 0.54 | 8.55 | 0.28 | |
| | Ethnicity | Mixed | 1.18 | 0.14 | 9.93 | 0.88 | |
| | Ethnicity | Asian | 1.63 | 0.61 | 4.38 | 0.34 | |
| | Ethnicity | Black | 1.72 | 0.54 | 5.46 | 0.36 | |
| | Ethnicity | Other | 0.76 | 0.16 | 3.59 | 0.73 | |
| | Ethnicity | Unknown | 3.54 | 0.71 | 17.63 | 0.12 | |
| | Number of body systems | Comorbidity present | 4.18 | 1.43 | 12.23 | 0.01 | |
| | Number of body systems | More than 1 body system | 5.56 | 2.16 | 14.30 | 0.00 | |

| | | | | | | | |
|---|---|---|---|---|---|---|---|
| Other pandemic year admission | Sex | Female | 0.59 | 0.50 | 0.68 | 0.00 | 111464 |
| | IMD | 2nd most deprived quintil | 1.24 | 0.99 | 1.54 | 0.06 | |
| | IMD | 3rd most deprived quintil | 1.05 | 0.83 | 1.33 | 0.68 | |
| | IMD | 4th most deprived quintil | 1.00 | 0.78 | 1.27 | 0.98 | |
| | IMD | Least deprived quintile | 1.42 | 1.12 | 1.79 | 0.00 | |
| | Ethnicity | Mixed | 1.44 | 0.97 | 2.15 | 0.07 | |
| | Ethnicity | Asian | 1.88 | 1.50 | 2.35 | 0.00 | |
| | Ethnicity | Black | 1.98 | 1.51 | 2.61 | 0.00 | |
| | Ethnicity | Other | 1.36 | 0.87 | 2.11 | 0.18 | |
| | Ethnicity | Unknown | 1.60 | 1.24 | 2.06 | 0.00 | |
| | Number of body systems | Comorbidity present | 11.23 | 6.05 | 20.84 | 0.00 | |
| | Number of body systems | More than 1 body system | 57.26 | 31.50 | 104.10 | 0.00 | |
| All admissions 2019/20 | Sex | Female | 0.56 | 0.49 | 0.64 | 0.00 | 147870 |
| | IMD | 2nd most deprived quintil | 0.91 | 0.75 | 1.12 | 0.38 | |
| | IMD | 3rd most deprived quintil | 0.97 | 0.79 | 1.20 | 0.80 | |
| | IMD | 4th most deprived quintil | 0.98 | 0.78 | 1.22 | 0.82 | |
| | IMD | Least deprived quintile | 1.07 | 0.86 | 1.34 | 0.55 | |
| | Ethnicity | Mixed | 1.03 | 0.69 | 1.54 | 0.89 | |
| | Ethnicity | Asian | 1.38 | 1.11 | 1.71 | 0.00 | |
| | Ethnicity | Black | 2.28 | 1.80 | 2.88 | 0.00 | |
| | Ethnicity | Other | 1.65 | 1.14 | 2.38 | 0.01 | |
| | Ethnicity | Unknown | 1.00 | 0.75 | 1.33 | 1.00 | |
| | Number of body systems | Comorbidity present | 7.27 | 4.43 | 11.93 | 0.00 | |
| | Number of body systems | More than 1 body system | 40.30 | 25.19 | 64.48 | 0.00 | |
| | Sex | Female | 1.67 | 0.76 | 3.64 | 0.20 | 654 |
| | IMD | 2nd most deprived quintil | 0.95 | 0.29 | 3.09 | 0.93 | |

| | | | | | | |
|---|---|---|---|---|---|---|
| | IMD | 3rd most deprived quintil | 1.28 | 0.44 | 3.78 | 0.65 |
| | IMD | 4th most deprived quintil | 1.04 | 0.34 | 3.13 | 0.95 |
| | IMD | Least deprived quintile | 0.70 | 0.13 | 3.65 | 0.67 |
| Influenza admissions 2019/20 | Ethnicity | Mixed | - | - | - | - |
| | Ethnicity | Asian | 1.51 | 0.55 | 4.11 | 0.43 |
| | Ethnicity | Black | 0.53 | 0.07 | 4.14 | 0.55 |
| | Ethnicity | Other | 1.99 | 0.21 | 18.86 | 0.55 |
| | Ethnicity | Unknown | 1.84 | 0.46 | 7.33 | 0.39 |
| | Number of body systems | Comorbidity present | 0.13 | 0.04 | 0.44 | 0.00 |
| | Number of body systems | More than 1 body system | - | - | - | - |

Data are odds ratios with confidence intervals for full multivariable GEE models, absolute predicted probability of PICU admission within each comorbidity group, and absolute difference in predicted probability of PICU admission compared with no comorbidity
Age groups are as per JCVI request - infants, 1-4 (baseline), 5-10, 11-15, 16-17
"Age group used " specifies if all age groups are included (JCVI) or results are stratified by secondary school CYP only (11-17)
Models are adjusted for sex, ethnicity, IMD and age (unless only 11-17 included). Estimates for full modesl are availbale in supplementary material 3.
